# Cefazolin versus Antistaphylococcal Penicillins for the Treatment of Methicillin-Susceptible *Staphylococcus aureus* Bacteremia: A Systematic Review and Meta-Analysis

**DOI:** 10.1101/2025.02.17.25322429

**Authors:** Connor Prosty, Dean Noutsios, Todd C. Lee, Nick Daneman, Joshua S. Davis, Nynke G. L. Jager, Nesrin Ghanem-Zoubi, Anna L. Goodman, Achim J. Kaasch, Ilse Kouijzer, Brendan J. McMullan, Emily G. McDonald, Steven Y. C. Tong, Sean W. X. Ong, the Staphylococcus aureus Network Adaptive Platform MSSA/PSSA domain specific working group

**Affiliations:** Faculty of Medicine, McGill University, Montréal, QC, Canada; Division of Experimental Medicine, Department of Medicine, McGill University, Montréal, QC, Canada; Division of Infectious Diseases, Department of Medicine, McGill University Health Centre, QC, Montréal, Canada; Clinical Practice Assessment Unit, Department of Medicine, McGill University Health Centre, Montréal, QC, Canada; Division of Infectious Diseases, Sunnybrook Health Sciences Centre, Toronto, ON, Canada; Institute of Health Policy, Management and Evaluation, University of Toronto, Toronto, ON, Canada; School of Medicine and Public Health, University of Newcastle, Newcastle, Australia; Department of Immunology and Infectious Diseases, John Hunter Hospital, Newcastle, Australia; Global and Tropical Health Division, Menzies School of Health and Research, Darwin, Australia; Department of Pharmacy, Radboud Institute for Medical Innovation, Radboud University Medical Center, Nijmegen, The Netherlands; Infectious Diseases Institute, Rambam Health Care Campus, Haifa, Israel; Medical Research Council Clinical Trials Unit, University College London, London, United Kingdom; Institute of Medical Microbiology and Hospital Hygiene, Faculty of Medicine, Heinrich Heine University Düsseldorf, Düsseldorf, Germany; Department of Internal Medicine and Radboud Center for Infectious Diseases, Radboud University Medical Center, Nijmegen, The Netherlands; Department of Infectious Diseases, Sydney Children’s Hospital, Randwick, New South Wales, Australia; Division of General Internal Medicine, Department of Medicine, McGill University Health Centre, Montréal, QC, Canada; Department of Infectious Diseases, The University of Melbourne at the Peter Doherty Institute for Infection and Immunity, Melbourne, Australia; Victorian Infectious Disease Service, The Royal Melbourne Hospital at the Peter Doherty Institute for Infection and Immunity, Melbourne, Australia

**Author notes:** **Corresponding author:** Connor Prosty, MD 1001 Decarie Blvd E5-1820, Montréal, Québec, H4A 3J1. **Funding:** This project received no funding.

**Keywords:** *Staphylococcus aureus*, bacteremia, blood stream infection, septicemia, cefazolin, antistaphylococcal penicillin

## Abstract

**Background:** There is debate on whether cefazolin or antistaphylococcal penicillins should be the first-line treatment for methicillin-susceptible *Staphylococcus aureus* (MSSA) bacteremia. Ongoing trials are investigating whether cefazolin is non-inferior to (flu)cloxacillin, but it remains uncertain whether these findings would apply to other antistaphylococcal penicillins.

**Objective:** We conducted a systematic review and meta-analysis comparing cefazolin to each of the individual antistaphylococcal penicillins for MSSA bacteremia.

**Methods:** *Data Sources:* We updated a 2019 systematic review but specifically focused on evaluating outcomes by individual antistaphylococcal penicillins.

*Study Eligibility Criteria:* Comparative observational studies.

*Participants:* Patients with MSSA bacteremia.

*Interventions:* Cefazolin versus the antistaphylococcal penicillins.

*Assessment of Risk of Bias:* The risk of bias in non-randomized studies of interventions tool.

*Methods of Data Synthesis:* The primary outcome was 30-day all-cause mortality, and we assessed for non-inferiority of cefazolin using a prespecified non-inferiority margin of a pooled odds ratio (OR) <1.2. Secondary outcomes were 90-day mortality, treatment-related adverse events (TRAEs), discontinuation due to toxicity, and nephrotoxicity.

**Results:** No randomized data have been published. 30 observational studies at moderate or high risk of bias were included, which comprised 3869 patients who received cefazolin and 11644 patients who received antistaphylococcal penicillins (flucloxacillin=6721, unspecified=2440, nafcillin=1305, cloxacillin=1258, and oxacillin=120). Cefazolin was associated with a reduced odds of 30-day all-cause mortality (OR=0.73, 95%CI=0.62-0.85) compared to antistaphylococcal penicillins, meeting pre-specified non-inferiority as well as superiority. This effect was consistent versus flucloxacillin (OR=0.92, 95%CI=0.73-1.16), nafcillin (OR=0.58, 95%CI=0.28-1.17), cloxacillin (OR=0.42, 95%CI=0.11-1.58), and oxacillin (OR=0.31, 95%CI=0.03-2.75). Point estimates favored cefazolin for 90-day mortality, TRAEs, nephrotoxicity, and discontinuation due to toxicity overall and in each comparison with individual antistaphylococcal penicillins, except for TRAEs versus cloxacillin.

**Conclusions:** In moderate to low quality observational data, cefazolin was associated with superior effectiveness and safety as compared to antistaphylococcal penicillins overall and individually.

## INTRODUCTION

*Staphylococcus aureus* bacteremia (SAB) is common, with an incidence ranging from 9-65 cases per 100,000 person-years,[1] and morbid with a 90-day all-cause mortality of ∼30%[2]. While there are multiple treatment options for methicillin-susceptible *S. aureus* (MSSA) bacteremia, there is no international consensus as to whether cefazolin or an antistaphylococcal penicillin is the optimal first-line treatment[3]. There are theoretical concerns that cefazolin may be suboptimal, particularly for infections with a high organism burden like endocarditis, because of the risk of an inoculum effect[4]. Because no international consensus guidelines on MSSA bacteremia have been published, recommendations are often extrapolated from native valve MSSA infective endocarditis (IE) guidelines. The European Society of Cardiology favors (flu)cloxacillin or cefazolin[5], the American Heart Association recommends nafcillin or oxacillin[6], the British Society for Antimicrobial Chemotherapy prefers flucloxacillin[7], whereas the WikiGuidelines have no preference between cefazolin, (flu)cloxacillin, nafcillin, or oxacillin[8] (**Supplemental Table 1**). Drug availability also differs by setting (*e.g.,* flucloxacillin and cloxacillin are not commercially available in the United States).

In contrast to the theoretical concerns over the inoculum effect, the observational evidence suggests that cefazolin is associated with less nephrotoxicity and similar effectiveness to antistaphylococcal penicillins[9]. Two ongoing randomized controlled trials (RCTs), the *S. aureus* Network Adaptive Platform (SNAP) trial (NCT05137119)[10] and the CloCeBa trial (NCT03248063)[11], will determine whether cefazolin is non-inferior to (flu)cloxacillin for the treatment of MSSA bacteremia. However, these trials will not compare cefazolin to other antistaphylococcal penicillins, specifically nafcillin and oxacillin, which are the preferred treatment for MSSA bacteremia in certain countries. The individual antistaphylococcal penicillins have distinct pharmacokinetic, pharmacodynamic, and *in vitro* properties, which could influence relative efficacy[12,13]. Further, there is observational data outside of MSSA bacteremia suggesting that the antistaphylococcal penicillins may have distinct safety profiles[14,15]. There are limited data directly comparing the different antistaphylococcal penicillins; therefore, it remains unclear whether the relative efficacy of cefazolin versus (flu)cloxacillin will be extrapolatable to comparisons between cefazolin and other antistaphylococcal penicillins. Prior systematic reviews on the topic did not stratify comparisons by individual antistaphylococcal penicillins[9,16–20] and since the latest systematic review[9], new studies have been published. We therefore aimed to update the previous systematic review[9] while specifically seeking to compare cefazolin to each antistaphylococcal penicillin separately to determine if effects are consistent across the entire antistaphylococcal penicillin class, which will be unanswered by the ongoing trials.

## METHODS

### Protocol

This study was conducted according to a pre-specified protocol registered on PROSPERO (CRD42024586270) and was conducted in accordance with guidance from PRISMA[21], MOOSE[22], and the Cochrane Handbook for Systematic Reviews of Interventions[23].

### Search Strategy

The search strategy of PubMed, the Web of Science, the Cochrane Library of Systematic Reviews, and clinicaltrials.gov from Weis et al.[9], initially conducted from database inception to June 26, 2018, was updated from June 26, 2018 to August 29, 2024 (**Supplemental Table 2**). No language restrictions were used. Studies in languages other than French or English were translated to English using Google Translate.

### Study Selection

The search results were imported into Covidence[24], de-duplicated, and screened by title and abstract by two independent reviewers (CP and DN). The full texts of relevant articles were retrieved and were screened in duplicate for fulfillment of the inclusion and exclusion criteria. The references lists of included articles and prior systematic reviews[9,16–20] were screened for additional studies relevant for inclusion. Disagreements throughout were resolved by consensus and if consensus could not be achieved a third reviewer (SWXO) adjudicated.

### Inclusion and Exclusion Criteria

RCTs or observational studies comparing cefazolin to any antistaphylococcal penicillins for MSSA bacteremia (regardless of its susceptibility to penicillin) were included. Case reports or series and conference abstracts were excluded. Studies comparing adjunctive therapies (*e.g.,* ertapenem) or not reporting on the primary or secondary outcomes were excluded. Antistaphylococcal penicillins were defined as any of the following: cloxacillin, flucloxacillin, dicloxacillin, oxacillin, or nafcillin.

### Quality Assessment

Study quality was evaluated in duplicate using the Cochrane Risk of Bias in Non-Randomized Studies I (ROBINS-I) tool[25]. Overall ratings corresponded to the lowest quality ranking in any one domain[25]. Disagreements were resolved by consensus. The *robvis* package[26] was used for visualization of quality assessments.

### Outcomes

The primary outcome was 30-day all-cause mortality for cefazolin versus the individual antistaphylococcal penicillins. For the primary outcome, we prespecified a non-inferiority margin of less than 1.2 for the upper bound of the 95% confidence interval (95%CI) of the odds ratio (OR) for cefazolin versus the antistaphylococcal penicillins, corresponding to the non-inferiority margin used for the SNAP trial[10]. The secondary outcomes were 90-day all-cause mortality, treatment-related adverse events (TRAE, as defined by the study), treatment discontinuation due to toxicity, and nephrotoxicity (as defined by the study). Analyses were conducted for cefazolin versus the antistaphylococcal penicillins, stratified by the individual antistaphylococcal penicillins. Subgroup effects by antistaphylococcal penicillins were tested by the Q-test[23]. A post hoc subgroup analysis was conducted for patients with IE.

### Data Extraction

In addition to the primary and secondary outcomes, the following variables were extracted in duplicate by independent reviewers: author, year, study design, number of centers, country, sex, study arms, and mean/median age, as well as the proportion admitted to the intensive care unit (ICU) and with IE at study entry. Adjusted data from propensity matched cohorts were extracted over unadjusted data, when presented. Otherwise, raw data were extracted. When data pertaining to the primary or secondary outcomes were unreported or if only pooled data across antistaphylococcal penicillins were reported instead of stratified by individual antistaphylococcal penicillins, study authors were contacted by email up to 3 times to request these data.

### Statistical Analyses

Analyses were performed with R (Version 4.3.2, R Foundation for Statistical Computing, Vienna, Austria) using the *metafor*[27] and *meta*[28] packages. Data were pooled by a frequentist random-effects inverse variance meta-analysis using a DerSimonian and Laird estimator for inter-study variance[29]. A 0.5 continuity correction was added for 0 event cells. I^2^ was used to evaluate inter-study heterogeneity; values >50% were considered as significant heterogeneity[23]. 95% confidence intervals (95%CI) were computed using a Wald-type normal distribution.

### Sensitivity Analyses

We pre-specified the following sensitivity analyses for the primary outcome: (i) restricting the analysis to studies at low or moderate risk of bias, (ii) including only studies conducted in high income countries, based on the World Bank classification (as drug availability and outcomes may differ significantly across high vs. low-income settings), and (iii) patients with IE. Publication bias of the primary outcome was assessed by visual inspection of a funnel plot and by Egger’s test[23]. The influence of individual studies on the overall estimate of the primary outcome was evaluated by a leave-one-out meta-analysis[23].

### Certainty of Evidence

The certainty of evidence for the primary and secondary outcomes was evaluated in duplicate by independent reviewers using the GRADE criteria[30] and presented using GRADEpro GDT[31].

## RESULTS

### Search Results and Population Characteristics

A total of 263 records were screened, 45 were evaluated by full text, of which, 15 were excluded with justification (**Figure 1**). The remaining 30 studies[32–61] were published from 2011-2024 and spanned 10 high-income countries. Four of the included studies were restricted to patients with IE[38,40,44,62], three focused on only penicillin-susceptible *S. aureus*[49,55,57], and one was limited to patients on hemodialysis[54]. A total of 3869 patients received cefazolin and 11644 received antistaphylococcal penicillins. Flucloxacillin was the most commonly administered antistaphylococcal penicillin (N=6721, 4 studies[37,42,55,56]), followed by nafcillin (N=1305, 13 studies[33,36,39,41,43,45,48,50,52,57,58,60,61]), cloxacillin (N=1258, 7 studies[34,38,40,49,51,54,59]), and oxacillin (N=120, 4 studies[41,46,53,60]). Data stratified by individual antistaphylococcal penicillin was not available for four studies[35,44,47,62] (N=2240) and 2 studies reported data on two antistaphylococcal penicillins[41,60]. The population was predominantly male (67.1%) and the mean/median ages ranged from 50-71 years. Further study details are presented in **Supplemental Tables 3 and 4**.

**Figure 1.**
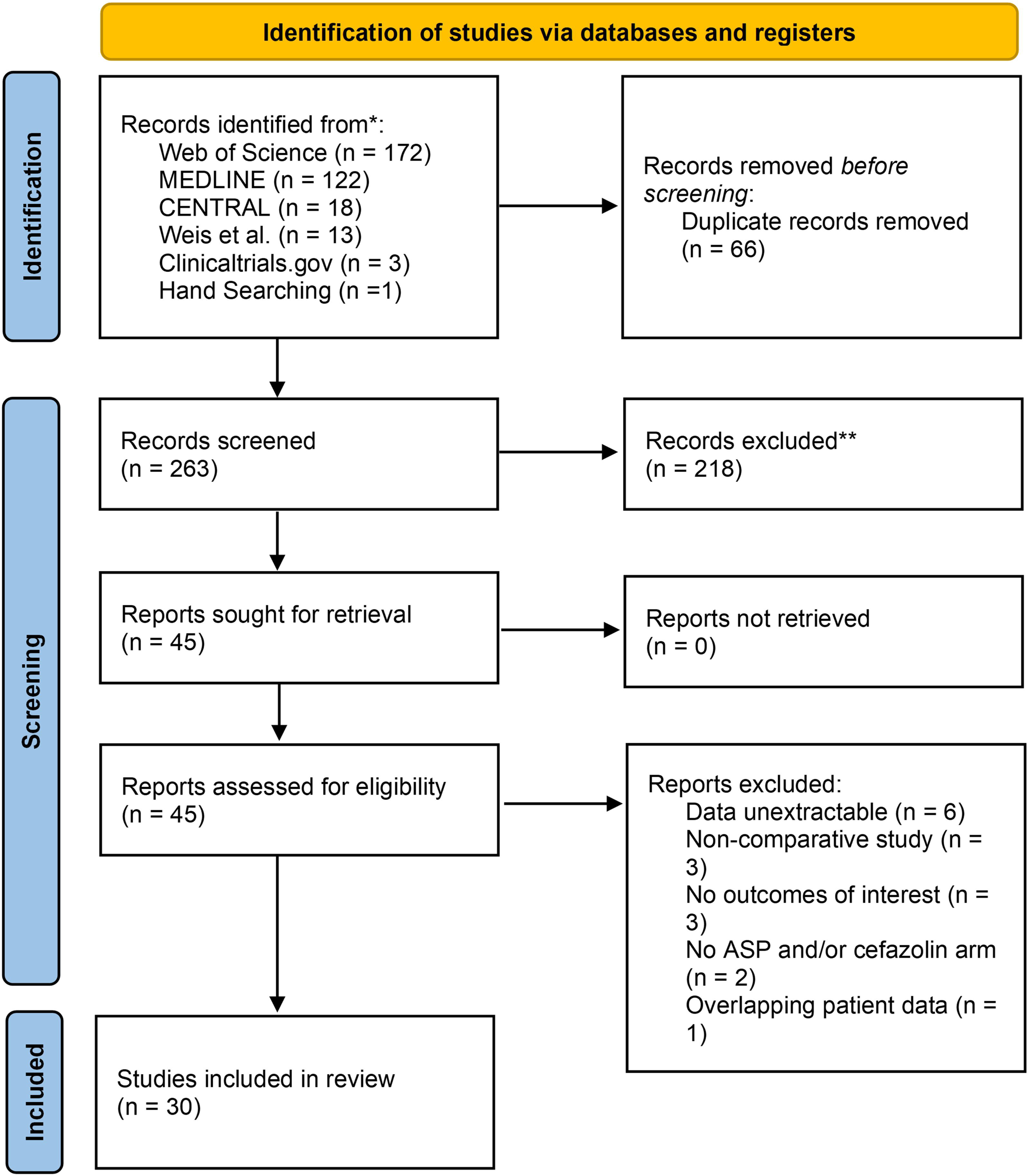
PRISMA diagram.

### Study Quality

The majority of studies (N=27, 90.0%) were at high risk of bias[32–34,36–42,44,46–61] due to concerns spanning multiple domains, and the remaining 3 studies (10.0%) were at moderate risk of bias[35,43,45] (**Figure 2**). First, adjustments for confounding by indication were inadequate or absent. Second, there was potential for immortal time bias, especially when patients were classified according to their definitive therapy. Third, important co-interventions such as surgical interventions or adjunctive agents were often imbalanced or unreported. Fourth, loss to follow-up was frequently unreported and no study had a pre-specified statistical analysis plan. Fifth, there was potential for ascertainment bias in the measurement of TRAEs and nephrotoxicity given that outcome assessment was not blinded.

**Figure 2.**
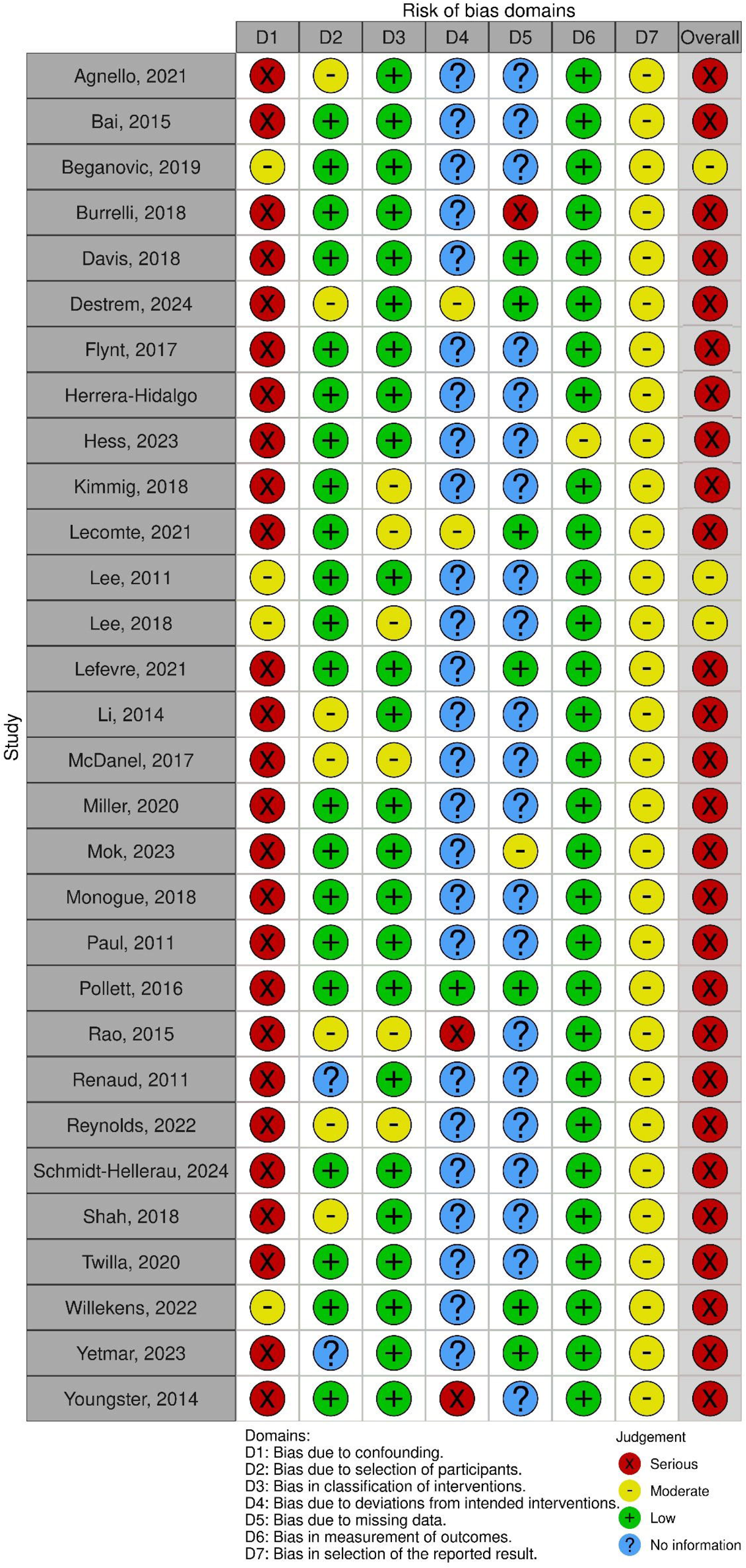
Risk of bias assessment.

### 30-Day All-Cause Mortality

In the 16 studies that reported on 30-day all-cause mortality, cefazolin was associated with a reduced odds of 30-day mortality compared to antistaphylococcal penicillins (8.6% [227/2631] vs. 11.9% [1113/9338], OR=0.73, 95%CI=0.62-0.85, I^2^=0.0%, 16 studies, **Figure 3**). There was no significant difference in treatment effect within a subgroup analysis of the individual antistaphylococcal penicillins (P=0.3). Point estimates continued to favor cefazolin when results were stratified by individual antistaphylococcal penicillins, but none were statistically significant, including: flucloxacillin (10.6% [92/865] vs. 11.3% [752/6665], OR=0.92, 95%CI=0.73-1.16, I^2^=0%, 3 studies), nafcillin (3.5% [16/462] vs. 6.8% [33/486], OR=0.58, 95%CI=0.28-1.17, I^2^=0.0%, 7 studies), cloxacillin (7.9% [3/38] vs. 18.6% [18/97], OR=0.42, 95%CI=0.11-1.58, I^2^=0.0%, 3 studies), and oxacillin (1.8% [4/227] vs. 2.3% [1/42], OR=0.31, 95%CI=0.03-2.75, I^2^=0.0%, 2 studies). Cefazolin met the pre-specified margin for non-inferiority when compared to antistaphylococcal penicillins as an entire group, as well as flucloxacillin and nafcillin individually, which had the largest number of patients available for comparisons.

**Figure 3.**
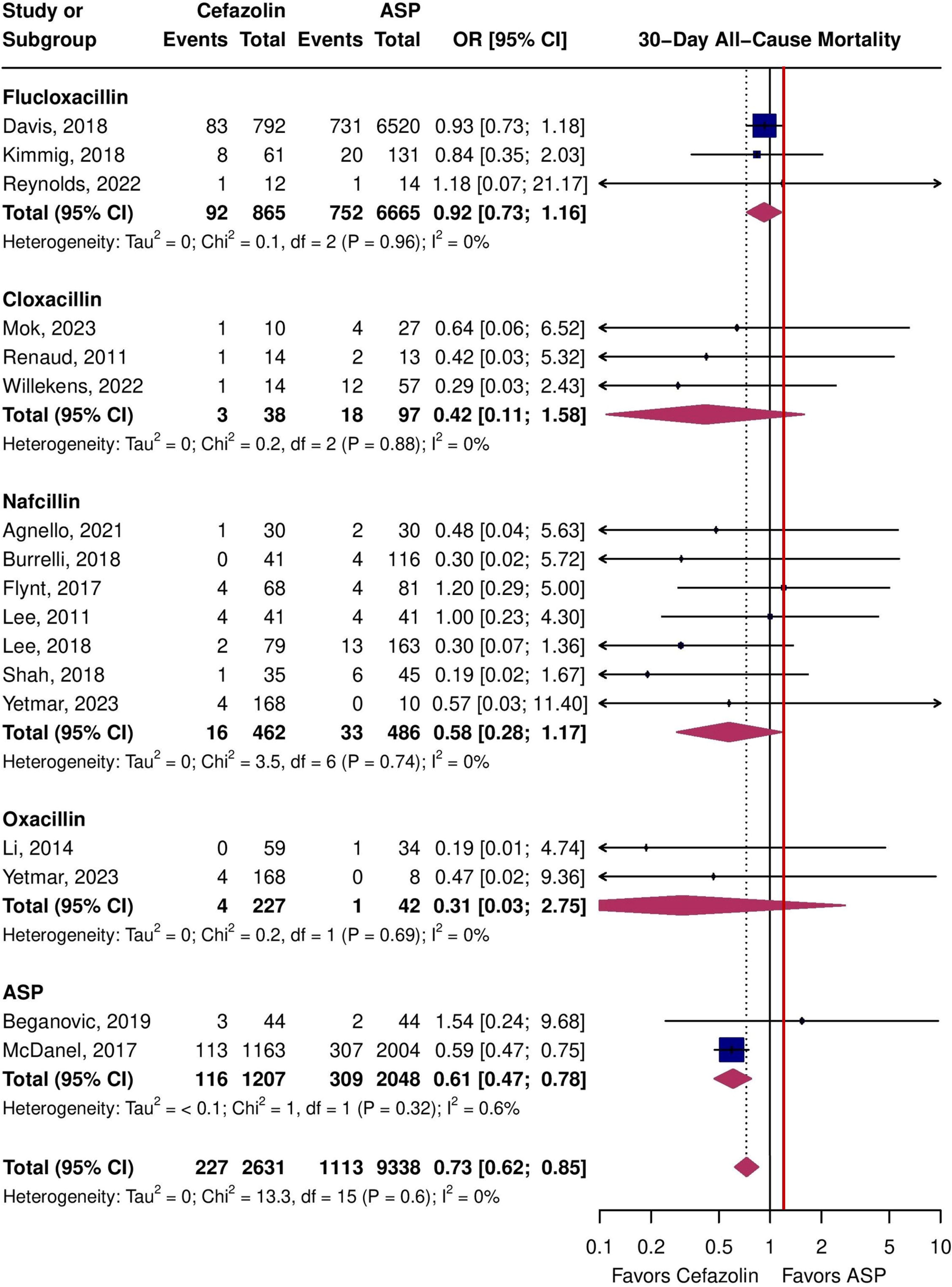
Forest plot of 30-day all-cause mortality for cefazolin vs. individual antistaphylococcal penicillins (ASP), the red line represents the non-inferiority margin.

### 90-Day All-Cause Mortality

The point estimate for 90-day all-cause mortality favored cefazolin and met the prespecified non-inferiority margin (17.1% [359/2097] vs. 24.1% [801/3318], OR=0.80, 95%CI=0.61-1.05, I^2^=29.0%, 14 studies, **Figure 4**). Effects were consistent across individual antistaphylococcal penicillins (P=0.8) and all favored cefazolin without achieving statistical significance.

**Figure 4.**
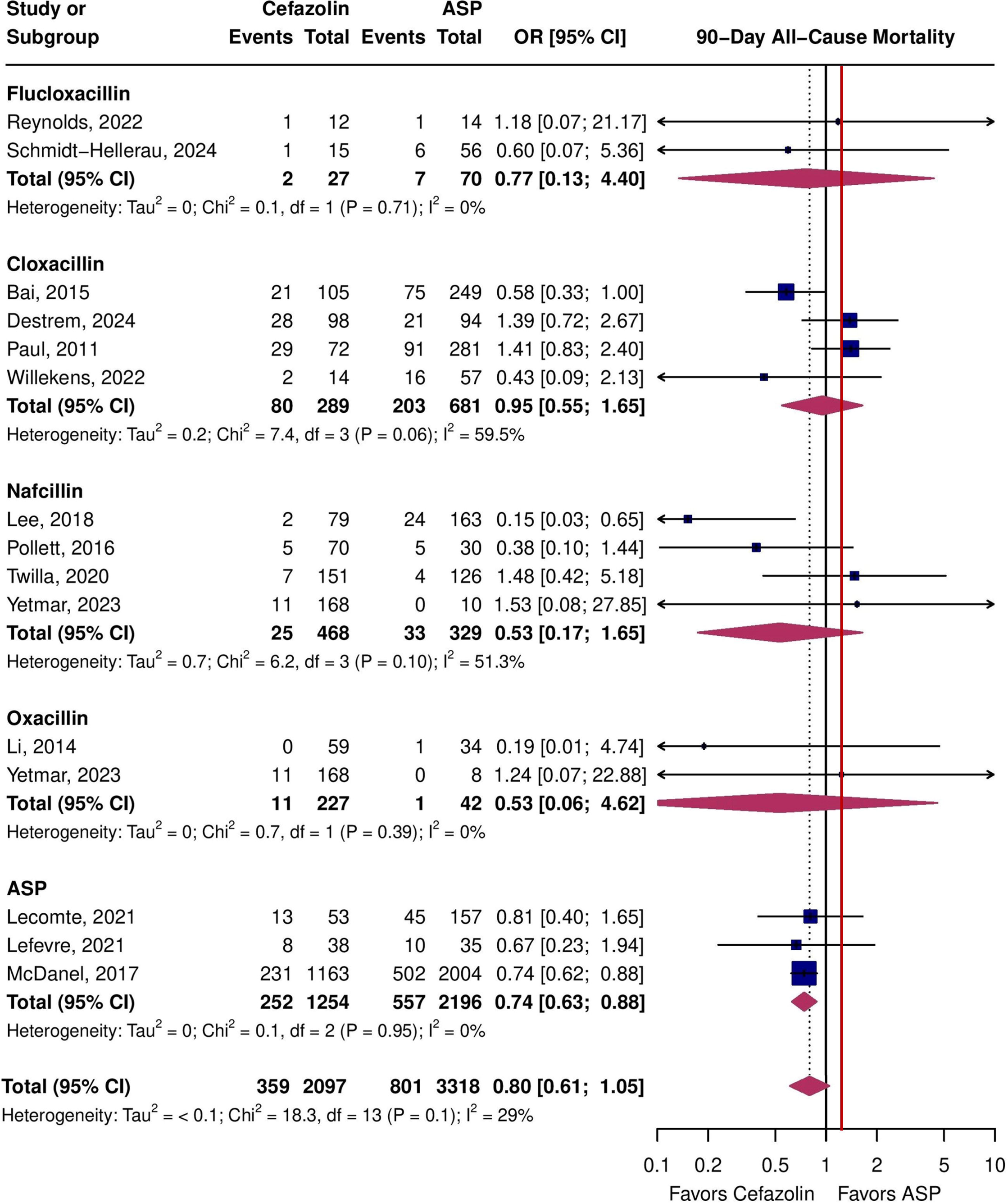
Forest plot of 90-day all-cause mortality for cefazolin vs. individual antistaphylococcal penicillins (ASP), the red line represents the non-inferiority margin.

### Safety Outcomes

Cefazolin was associated with a reduced odds of TRAEs compared to antistaphylococcal penicillins (11.0% [116/1052] vs. 23.5% [412/1755], OR=0.33, 95%CI=0.18-0.63, I^2^=75.8%, 14 studies, **Figure 5**). There were significant imbalances in subgroup analyses by individual antistaphylococcal penicillins (P<0.1). The point estimate favored cloxacillin (7.4% [6/81] vs. 2.4%, OR=1.65, 95%CI=0.47-5.87, I^2^=0.0%, 3 studies). However the remaining estimates favored cefazolin over nafcillin (8.7% [67/771] vs. 35.3% [364/1031], OR=0.53, 95%CI=0.17-1.65, I^2^=53.2%, 8 studies) and oxacillin (2.8% [11/397] vs. 10.7% [12/112], OR=0.39, 95%CI=0.04-4.22, I^2^=76.6%, 3 studies). No studies on flucloxacillin reported TRAEs.

**Figure 5.**
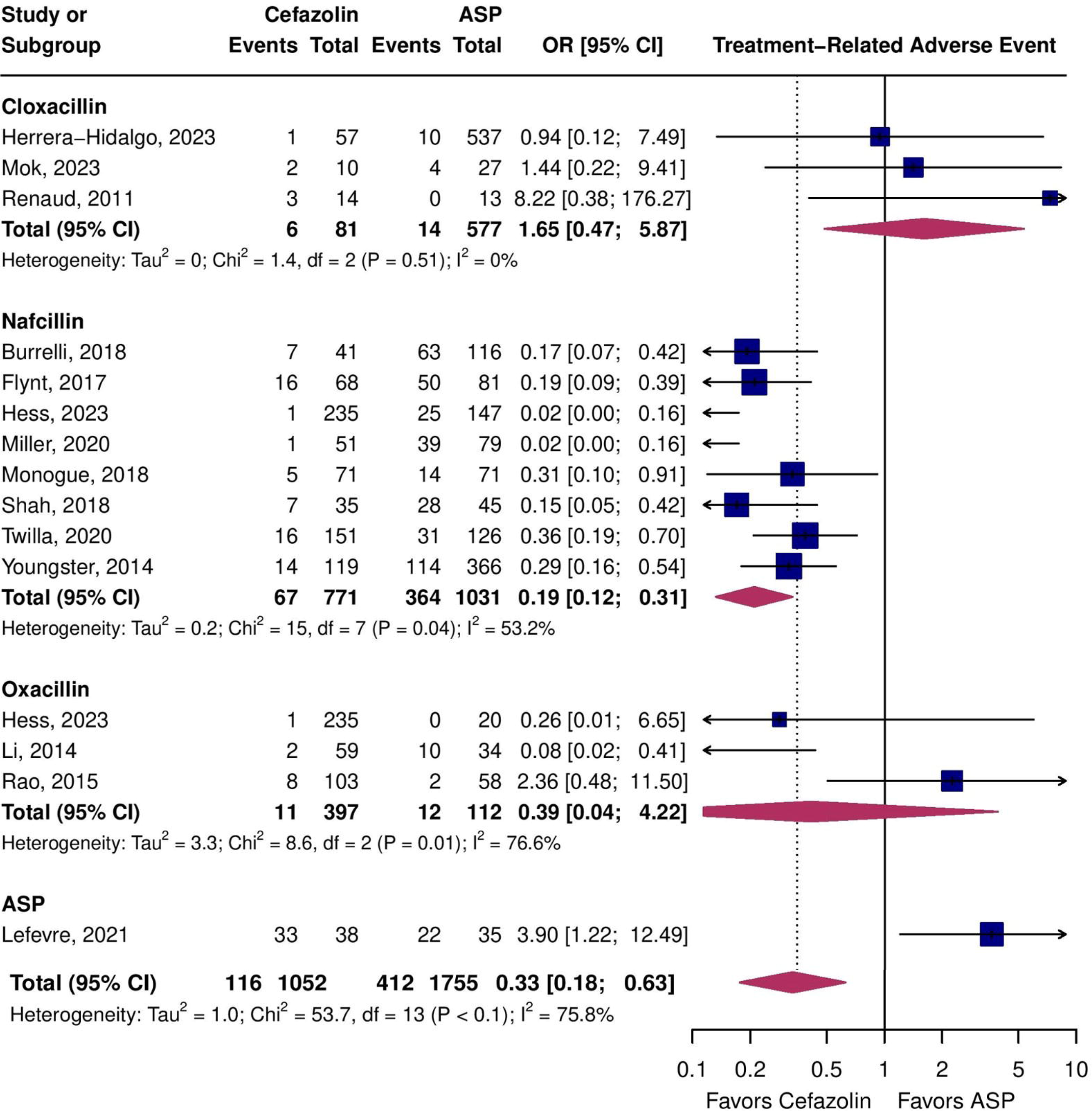
Forest plot of treatment-related mortality for cefazolin vs. individual antistaphylococcal penicillins (ASP).

Discontinuation due to toxicity was reduced with cefazolin compared to antistaphylococcal penicillins (2.1% [14/669] vs. 16.8% [142/843], OR=0.13, 95%CI=0.06-0.27, I^2^=20.4%, 10 studies, **Figure 6**) and effects were consistent across individual antistaphylococcal penicillins (P=0.8), with each estimate favoring cefazolin. Cefazolin was associated with significantly less toxicity than nafcillin (2.2% [12/542] vs. 20.0% [115/576], OR=0.09, 95%CI=0.03-0.30, I^2^=51.4%, 7 studies) and oxacillin (1.0% [3/294] vs. 13.0% [7/54], OR=0.15, 95%CI=0.04-0.67, I^2^=0.0%, 2 studies). There were insufficient studies to perform a meta-analysis of discontinuation due to toxicity for cloxacillin and flucloxacillin.

**Figure 6.**
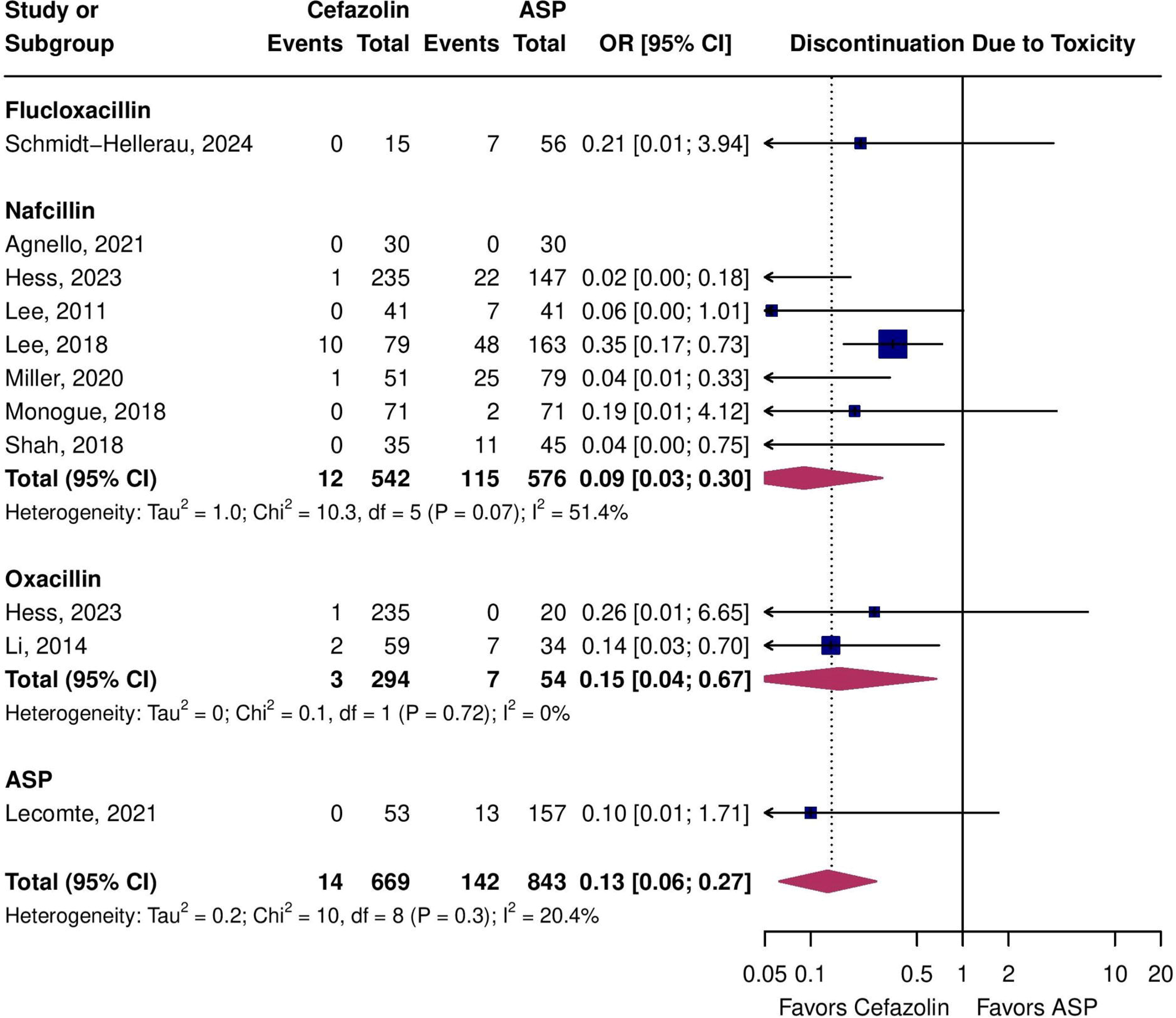
Forest plot of discontinuation due to toxicity for cefazolin vs. individual antistaphylococcal penicillins (ASP).

Nephrotoxicity was also lower with cefazolin versus antistaphylococcal penicillins (4.4% [48/1103] vs. 13.7% [205/1498], OR=0.30, 95%CI=0.20-0.46, I^2^=13.6%, 13 studies, **Figure 7**) and this effect was consistent across individual antistaphylococcal penicillins (P=0.5) with each estimate favoring cefazolin. Cefazolin was associated with significantly reduced nephrotoxicity compared to nafcillin (3.5% [30/850] vs. 15.2% [181/1194], OR=0.24, 95%CI=0.16-0.37, I^2^=0.0%, 9 studies), but not compared to oxacillin (0.3% [1/397] vs. 0.9% [1/112], OR=0.57, 95%CI=0.06-5.56, I^2^=0.0%, 3 studies). None of the flucloxacillin or cloxacillin studies reported of nephrotoxicity.

**Figure 7.**
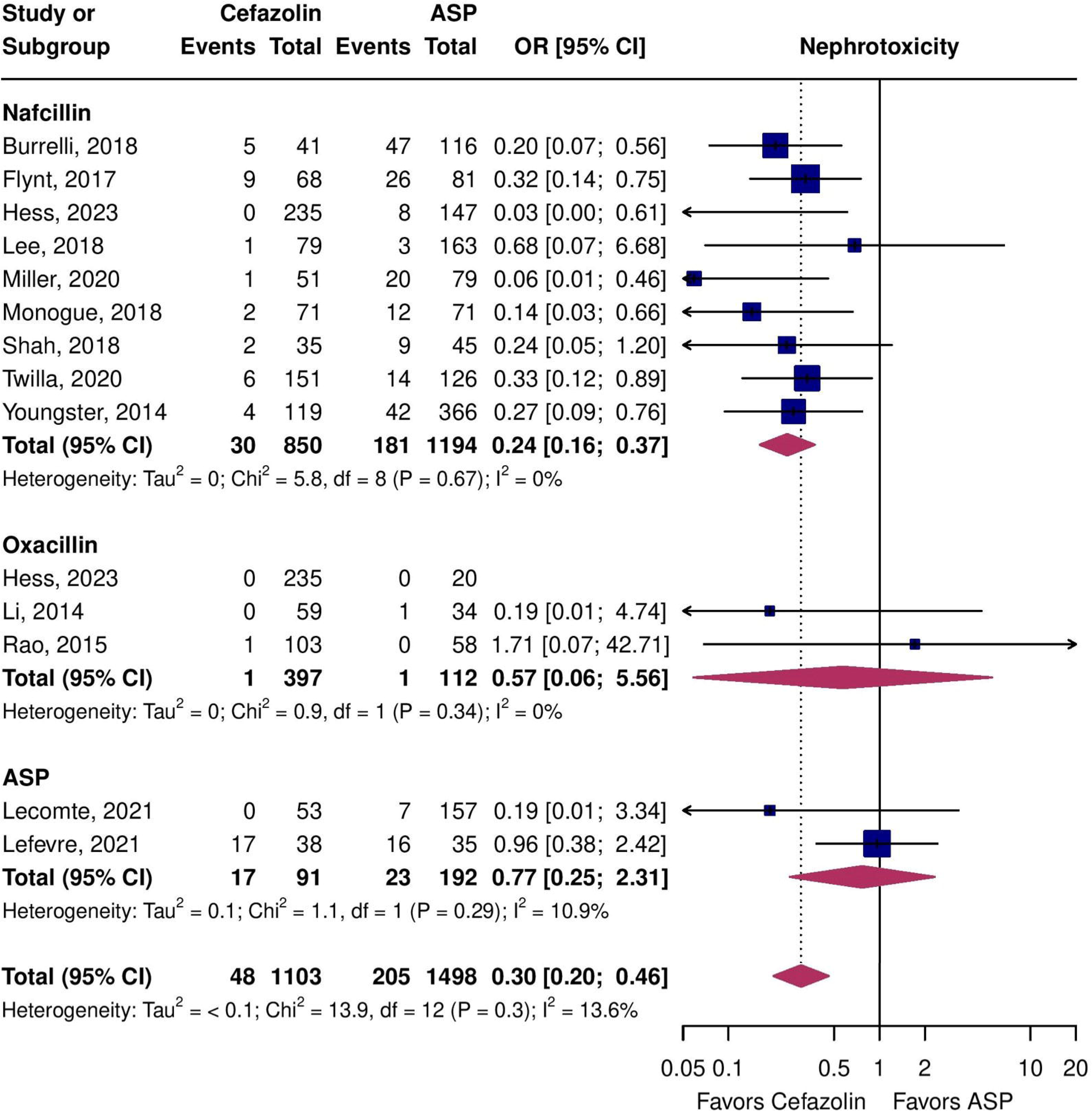
Forest plot of nephrotoxicity for cefazolin vs. individual antistaphylococcal penicillins (ASP).

### GRADE Recommendations

The certainty of evidence that, compared to antistaphylococcal penicillins, cefazolin was non-inferior for 30-day and 90-day all-cause mortality was low and very low, respectively. Certainty that cefazolin reduced: TRAEs was low; discontinuation due to toxicity was high; and nephrotoxicity was moderate (**Supplemental Table 5**).

### Subgroup Analyses

All studies were conducted in high income countries, which precluded this subgroup analysis. Excluding the studies at serious risk of bias favored cefazolin for 30-day all-cause mortality but was not statistically significant and did not meet the prespecified non-inferiority margin due to sample size (5.5% [9/164] vs. 7.7% [19/248], OR=0.72, 95%CI=0.28-1.85, I^2^=6.3%, 3 studies, **Supplemental Figure 1**). Subgroup analysis of patients with IE did not meet non-inferiority for 30-day all-cause mortality but cefazolin was favored (8.2% [5/61] vs. 15.7% [83/527], OR=0.35, 95%CI=0.05-2.72, I2=42.0%, 2 studies, **Supplemental Figure 2**).

There was no evidence of publication bias by funnel plot asymmetry or Egger’s test for the primary and secondary outcomes (**Supplemental Figures 3-7**). When the McDanel et al. study[47] was omitted from the meta-analysis of 30-day all-cause mortality, the pooled estimate was no longer statistically significant for superiority (OR=0.87, 95%CI=0.70-1.07, **Supplemental Figure 8**), but continued to meet non-inferiority. Otherwise, no studies with disproportionate impact on the overall estimates were identified (**Supplemental Figures 8-12**). Meta-regression was not performed because there were less than 10 studies[23] reporting each of the predictors of interest for the primary outcome.

## DISCUSSION

In this systematic review and meta-analysis of moderate to low quality observational studies, cefazolin was non-inferior to antistaphylococcal penicillins for 30-day mortality, non-inferior for 90-day mortality, and was associated with less toxicity across all safety outcomes. These results are consistent with previous reviews on the topic^8,11–15^; however, our study adds an additional 16 studies comprising 2654 patients compared to the most recent review[9].

Cefazolin was favorable compared to antistaphylococcal penicillins overall and individually (except cloxacillin for TRAEs) across all safety outcomes tested, including a 7-fold reduction in discontinuation due to toxicity and 3-fold reduction in occurrence of nephrotoxicity. This is an especially important consideration in the predominantly elderly and comorbid population with MSSA bacteremia, who frequently have chronic kidney disease[47] and for deep-seated MSSA infections like IE or osteomyelitis where therapy can be 6 weeks in duration. Cefazolin is also advantageous over antistaphylococcal penicillins because of less frequent dosing, which both reduces nursing resources needed for inpatients and facilitates outpatient parenteral antibiotic therapy. Cefazolin can be given thrice weekly on hemodialysis which eliminates the need for additional catheters in this population[63].

There are considerable geographic differences in the choice of antistaphylococcal penicillins or cefazolin as first-line therapy for MSSA bacteremia[3]. There is also regional variability in the selection and availability of antistaphylococcal penicillins that is reflected by differing international guideline recommendations[5–8] (**Supplemental Table 1**). The SNAP[10] and CloCeBa[11] trials are comparing cefazolin to (flu)cloxacillin. If these trials demonstrate that cefazolin is non-inferior to (flu)cloxacillin for mortality, particularly if associated with a superior safety profile, the results of this meta-analysis suggest that it would be reasonable to extrapolate the findings of SNAP and CloCeBa to the other antistaphylococcal penicillins (*i.e.,* nafcillin and oxacillin).

Strengths of this systematic review include a thorough exploration of multiple effectiveness and safety outcomes and stratification by individual antistaphylococcal penicillins. Further, the minimal statistical heterogeneity and consistent results across subgroups and sensitivity analyses bolster our findings. However, this review is subject to certain limitations, many of which are inherent to the studies included. First, most of the studies were at high risk of bias due to confounding by indication and the potential for immortal time bias. Second, between study differences in the definition and measurement of TRAEs and nephrotoxicity may produce heterogeneity. Third, we were unable to complete several pre-specified subgroup and sensitivity analyses due to a lack of granularity in the reported data. Fourth, there were no eligible studies of dicloxacillin, few studies of oxacillin, and pediatric data were lacking. Nevertheless, we believe that these results represent the most current and comprehensive comparison of cefazolin vs. antistaphylococcal penicillins.

## CONCLUSION

The current body of observational evidence suggests that cefazolin may be associated with reduced mortality and toxicity compared to antistaphylococcal penicillins. These results were consistent across comparisons with individual antistaphylococcal penicillins, suggesting that any differences compared to cefazolin are likely similar across the antistaphylococcal penicillins class.

## Data Availability

All data produced in the present study are available upon reasonable request to the authors

## ACKNOWLEDGEMENTS

The authors have no acknowledgements.

## AUTHOR CONTRIBUTIONS

Conceptualization - SWXO, CP, TCL, EGM, SYCT. Methodology - SWXO, CP, TCL, EGM, SYCT, DN. Software - CP, SWXO. Validation - CP, DN, SWXO. Formal Analysis - CP, SWXO. Investigation - CP, DN, SWXO. Resources - SWXO, TCL, EGM, SYCT. Data Curation - CP, DN. Writing Original Draft - All authors. Writing Review and Editing - All authors. Visualization - CP, SWXO. Supervision - SWXO, TCL, EGM, SYCT. Project administration - CP, SWXO, TCL, EGM, SYCT.

## SUPPLEMENTAL FIGURES

**Supplemental Figure 1.**
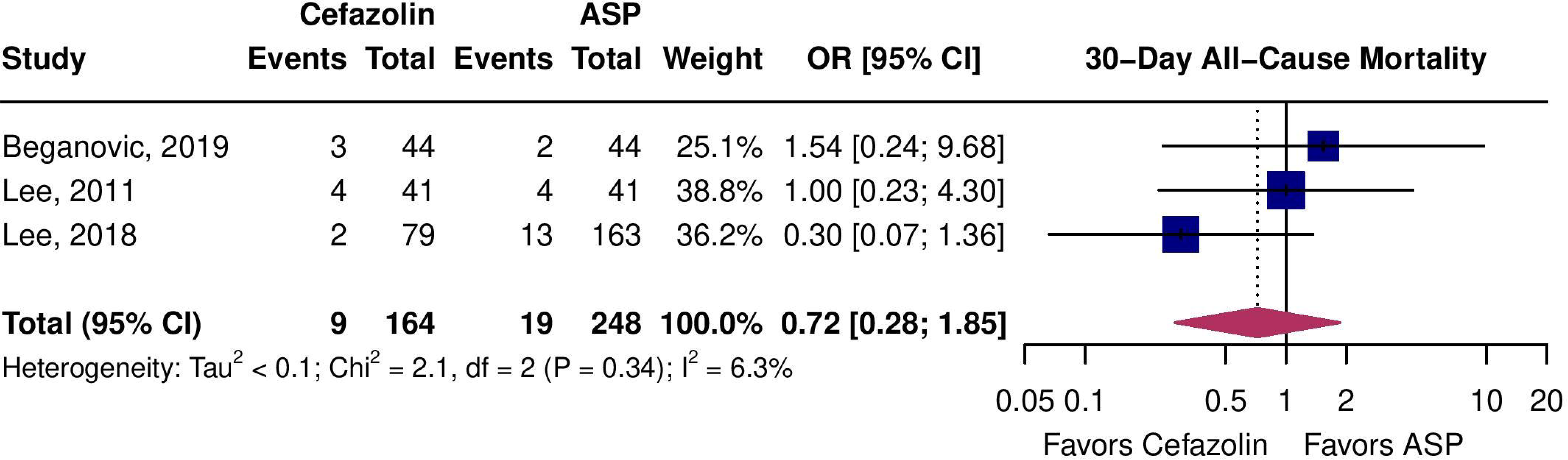
Forest plot of 30-day all-cause mortality for cefazolin vs. antistaphylococcal penicillins (ASP) excluding studies at high risk of bias.

**Supplemental Figure 2.**
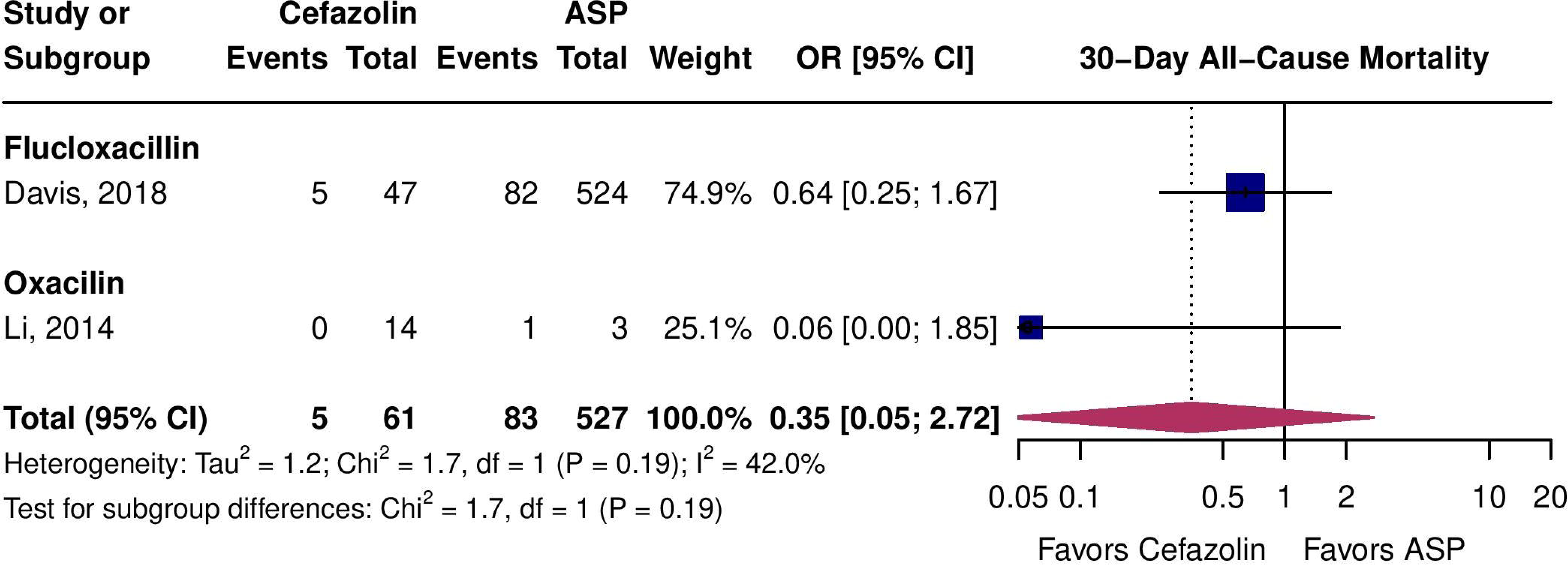
Forest plot of 30-day all-cause mortality for cefazolin vs. antistaphylococcal penicillins (ASP) in patients with IE.

**Supplemental Figure 3.**
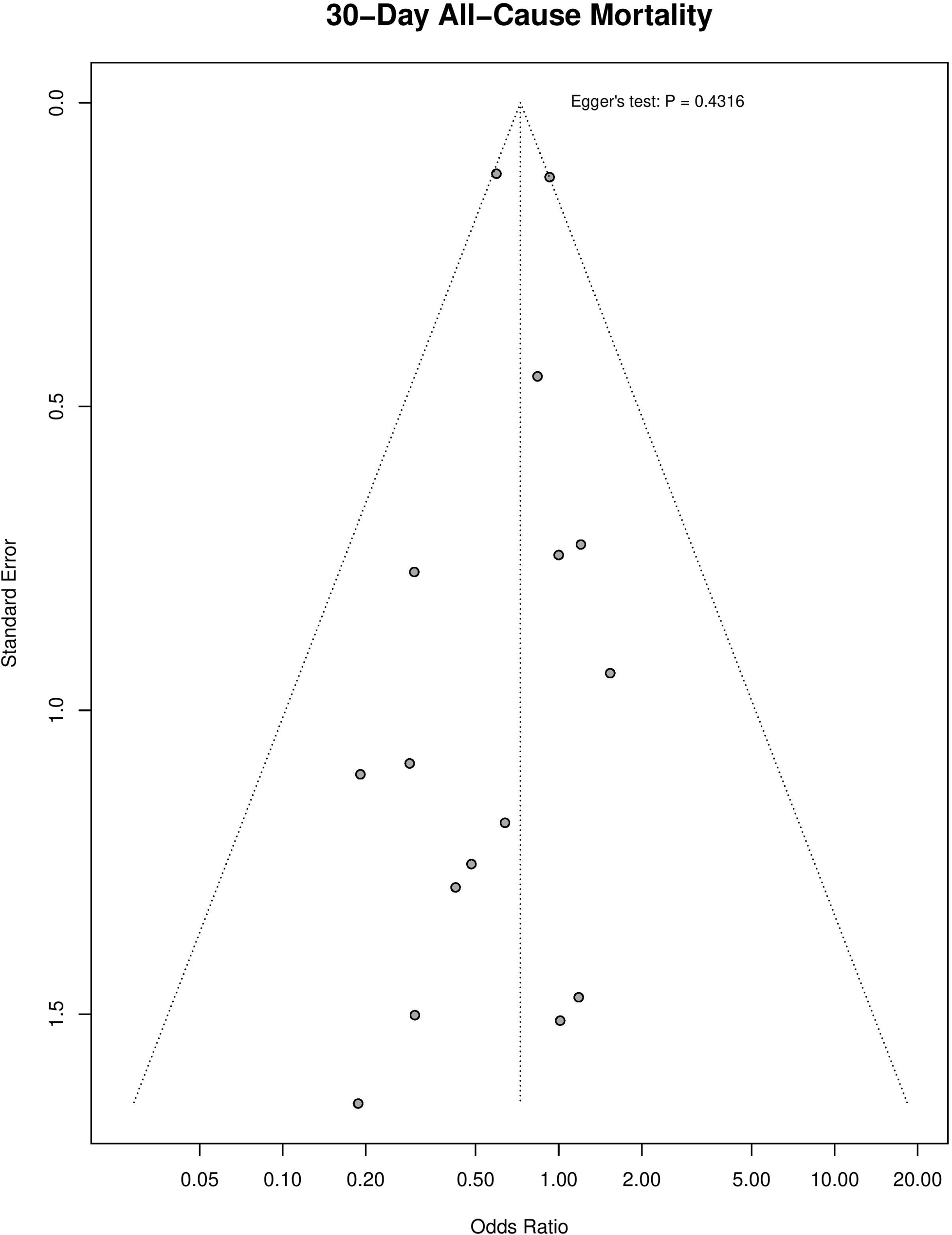
Funnel plot of 30-day all-cause mortality for cefazolin vs. antistaphylococcal penicillins (ASP).

**Supplemental Figure 4.**
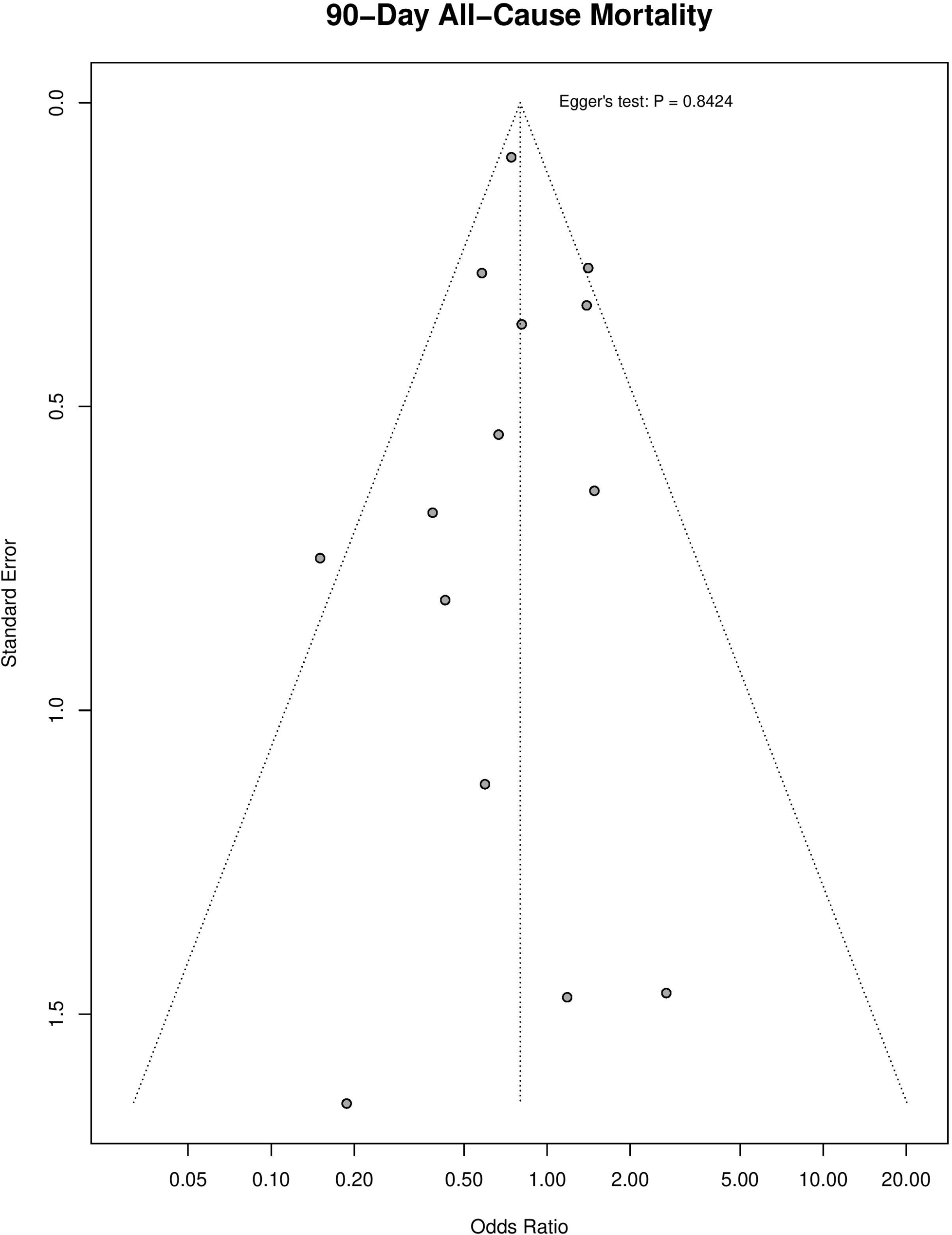
Funnel plot of 90-day all-cause mortality for cefazolin vs. antistaphylococcal penicillins.

**Supplemental Figure 5.**
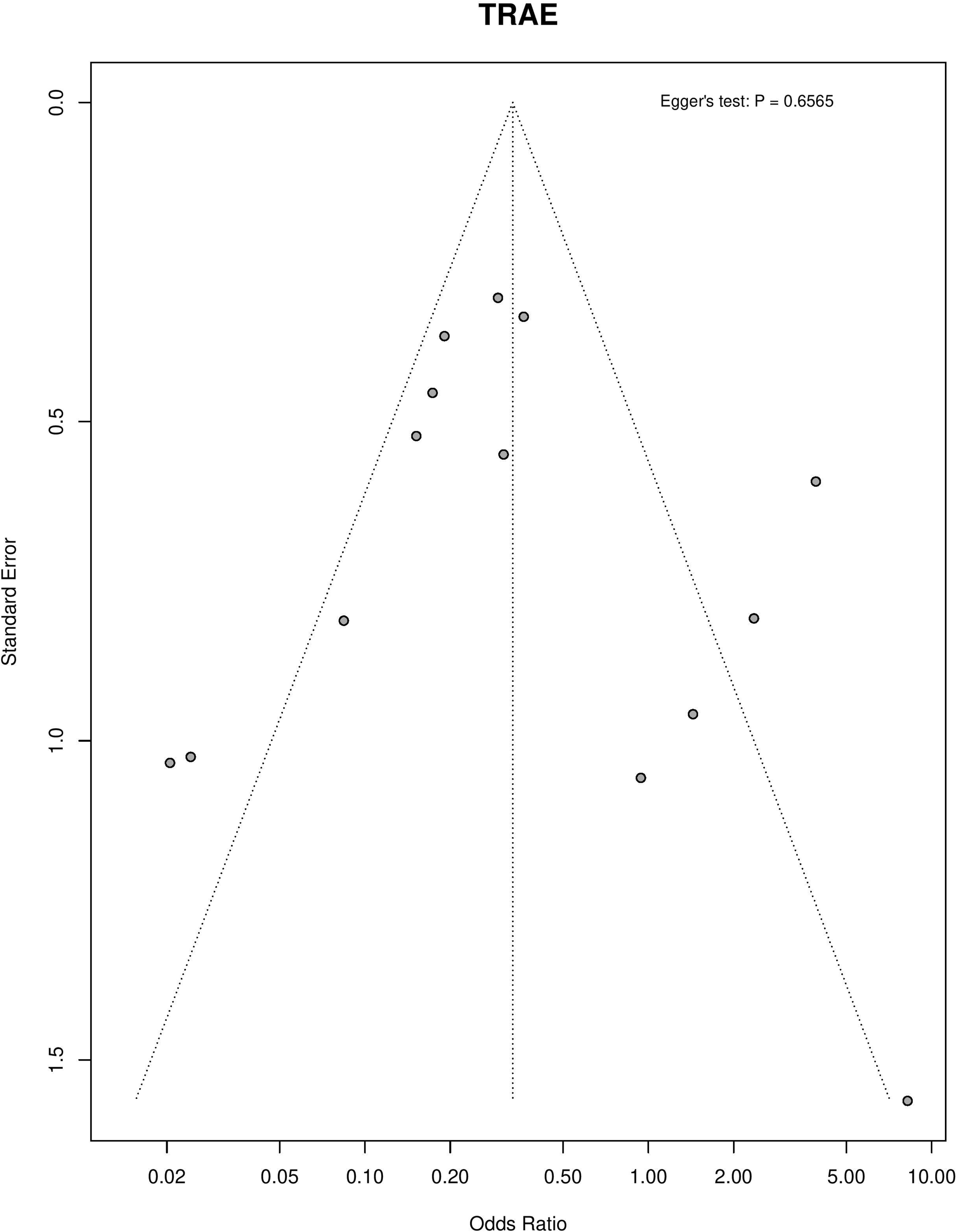
Funnel plot of treatment-related adverse events for cefazolin vs. antistaphylococcal penicillins.

**Supplemental Figure 6.**
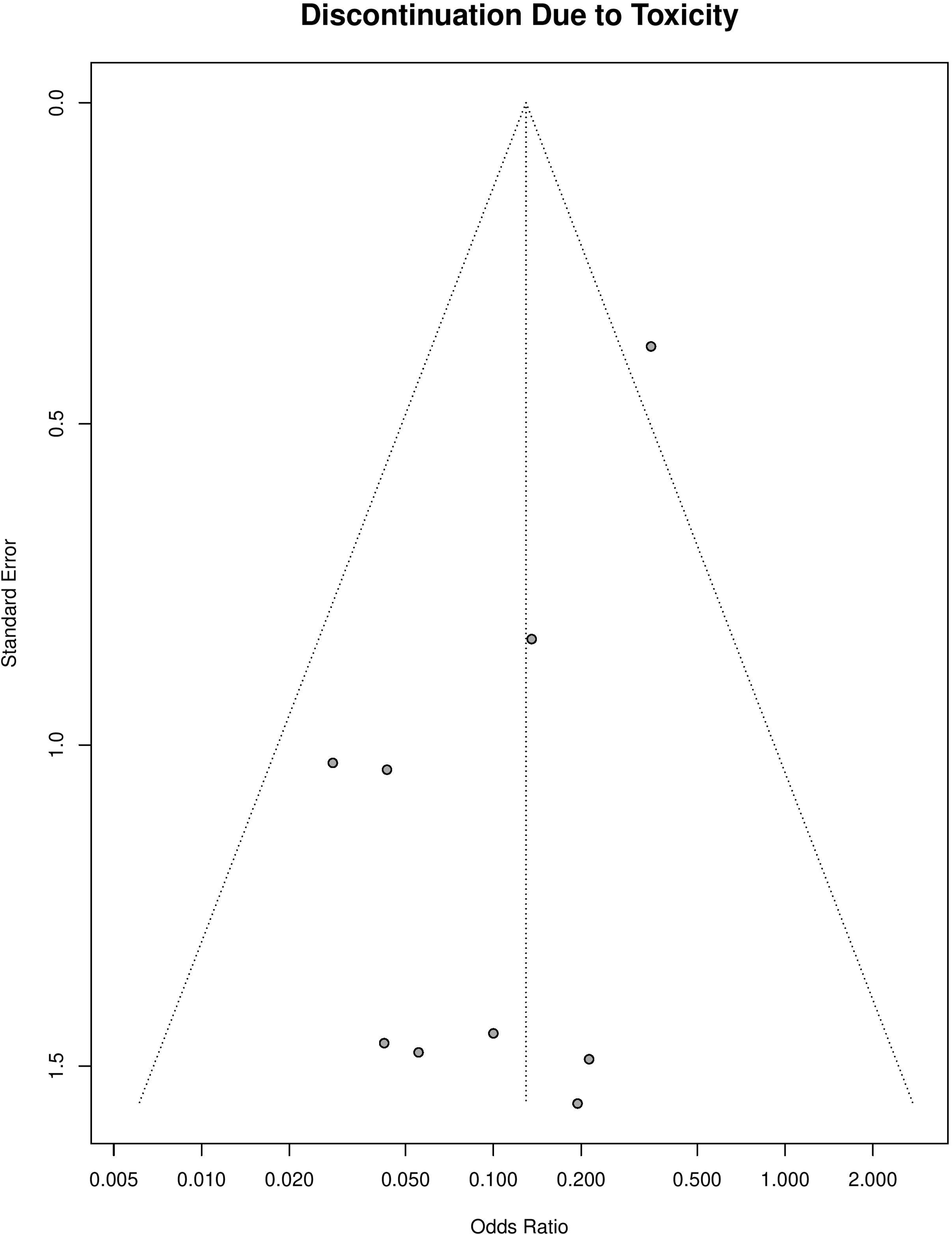
Funnel plot of discontinuation due to toxicity for cefazolin vs. antistaphylococcal penicillins. Egger’s test was not performed because there were <10 studies reporting outcome.

**Supplemental Figure 7.**
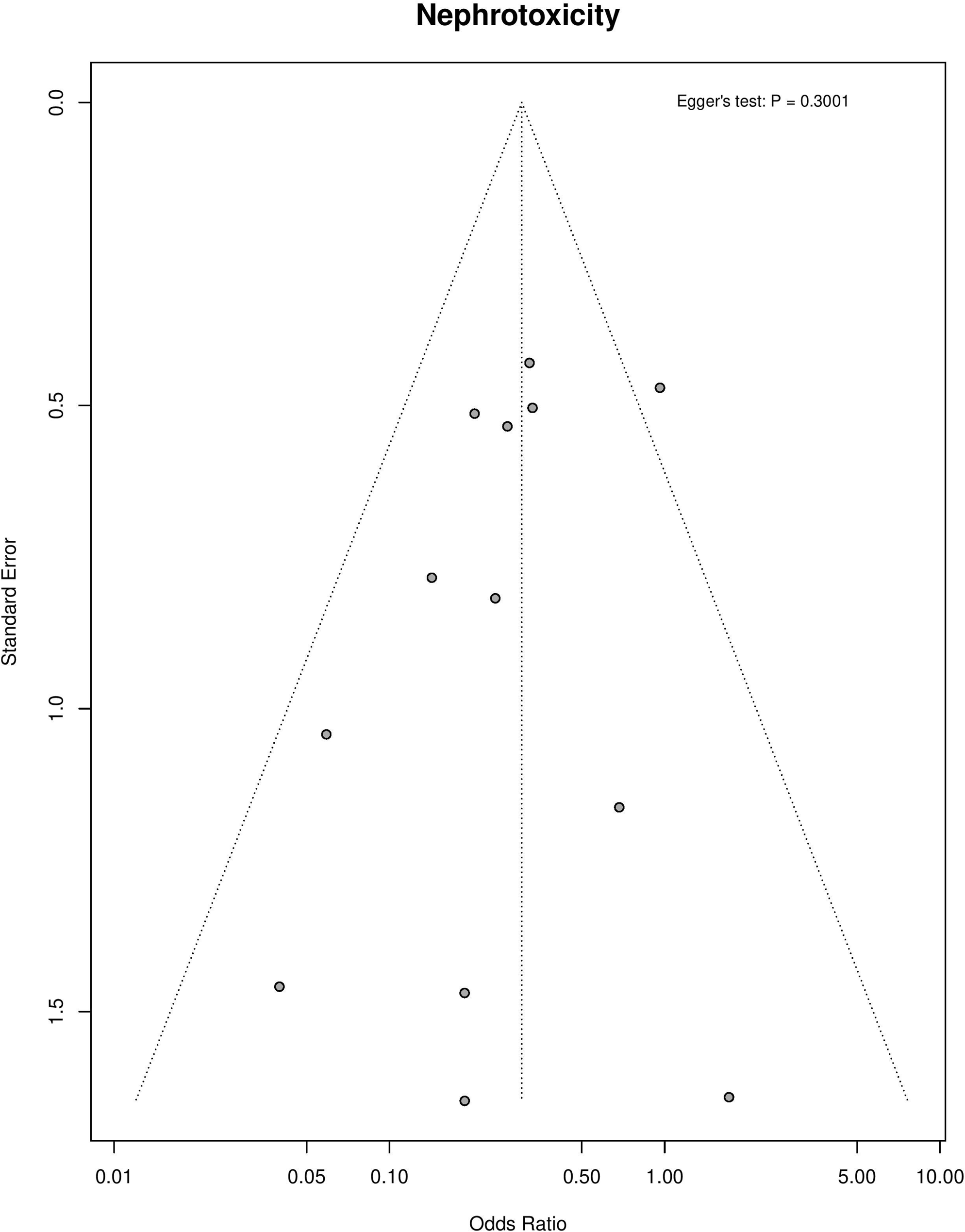
Funnel plot of nephrotoxicity for cefazolin vs. antistaphylococcal penicillins.

**Supplemental Figure 8.**
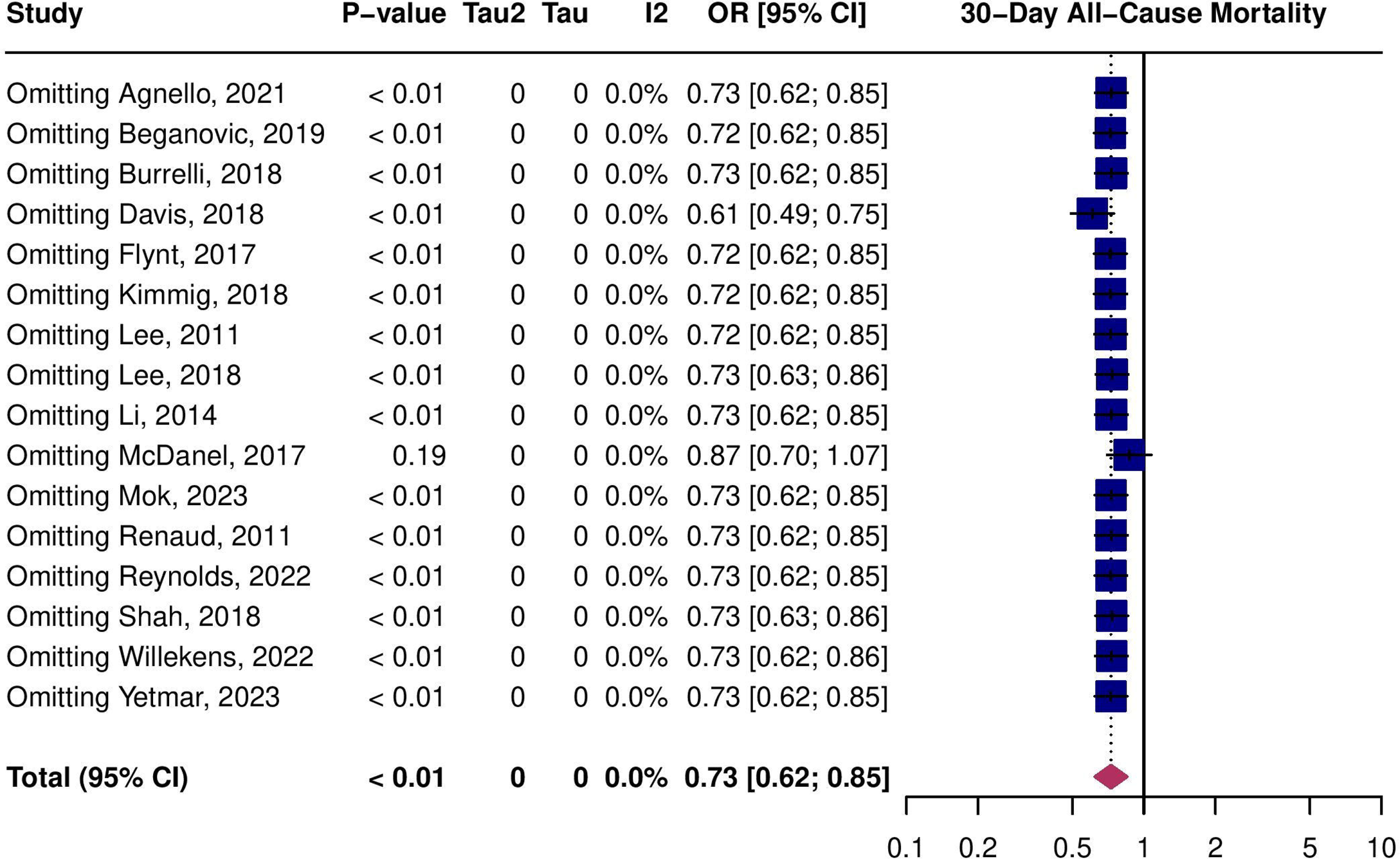
Leave-one-out meta-analysis of 30-day all-cause mortality for cefazolin vs. antistaphylococcal penicillins.

**Supplemental Figure 9.**
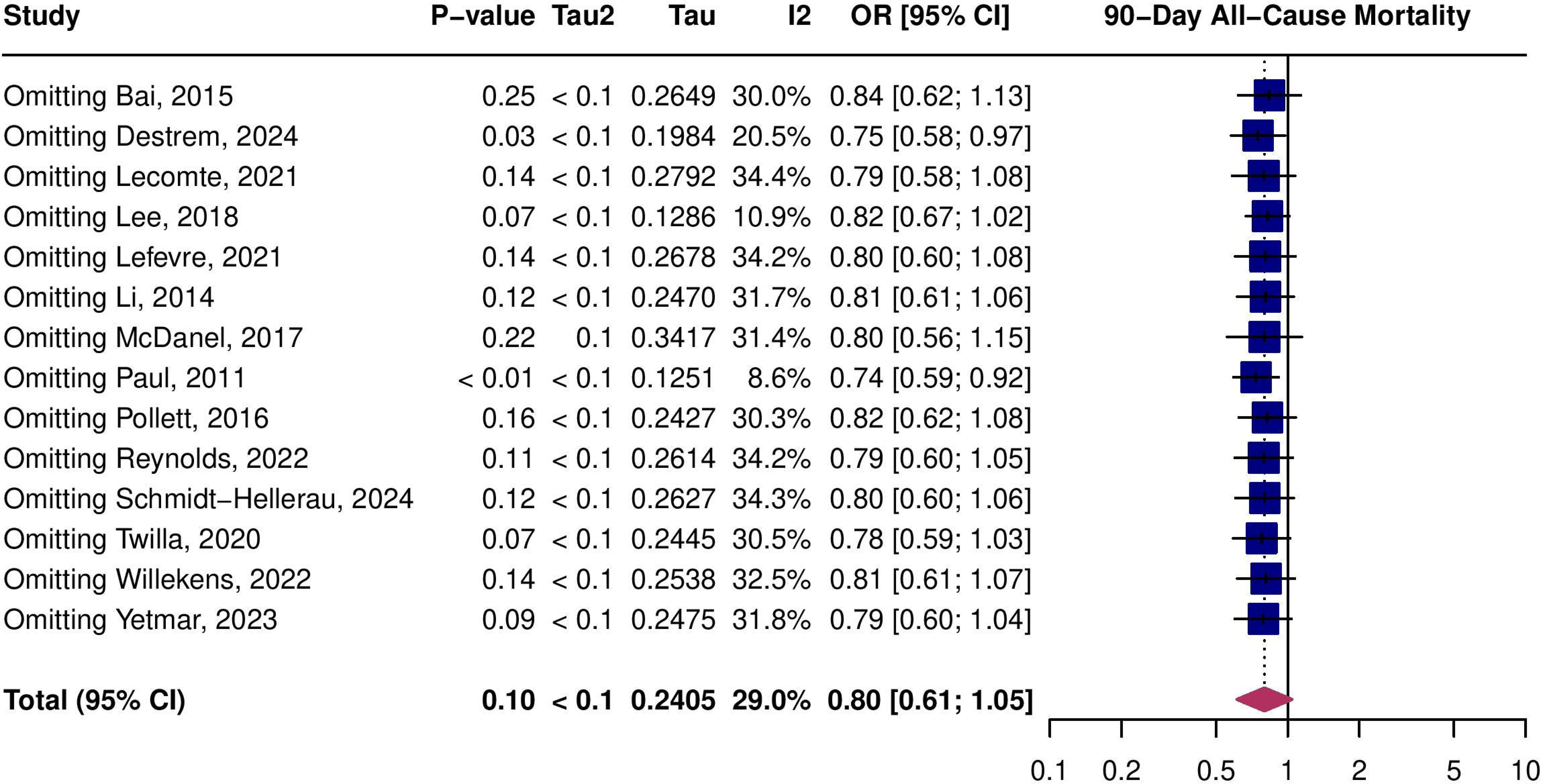
Leave-one-out meta-analysis of 90-day all-cause mortality for cefazolin vs. antistaphylococcal penicillins.

**Supplemental Figure 10.**
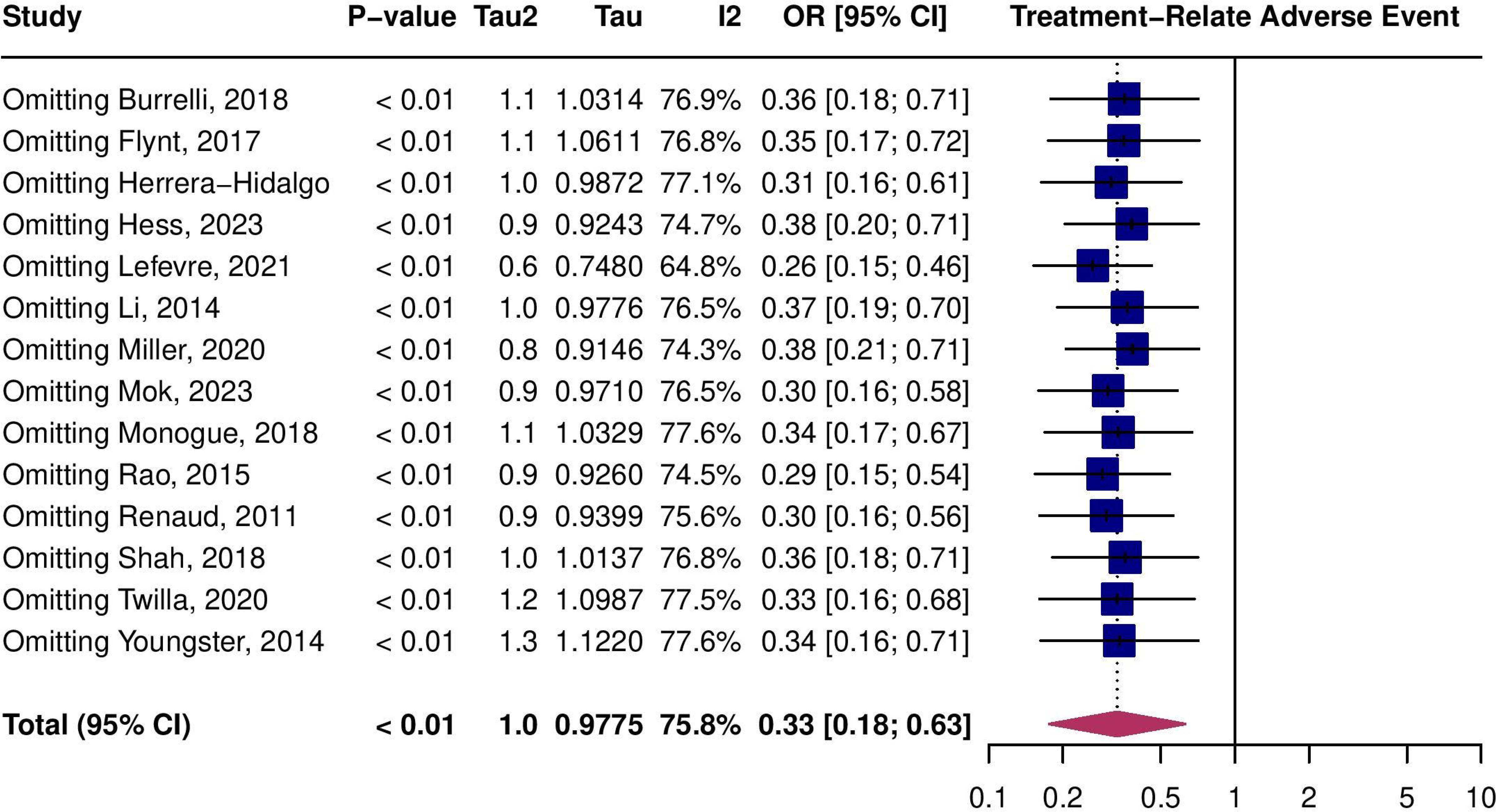
Leave-one-out meta-analysis of treatment-related adverse events for cefazolin vs. antistaphylococcal penicillins.

**Supplemental Figure 11.**
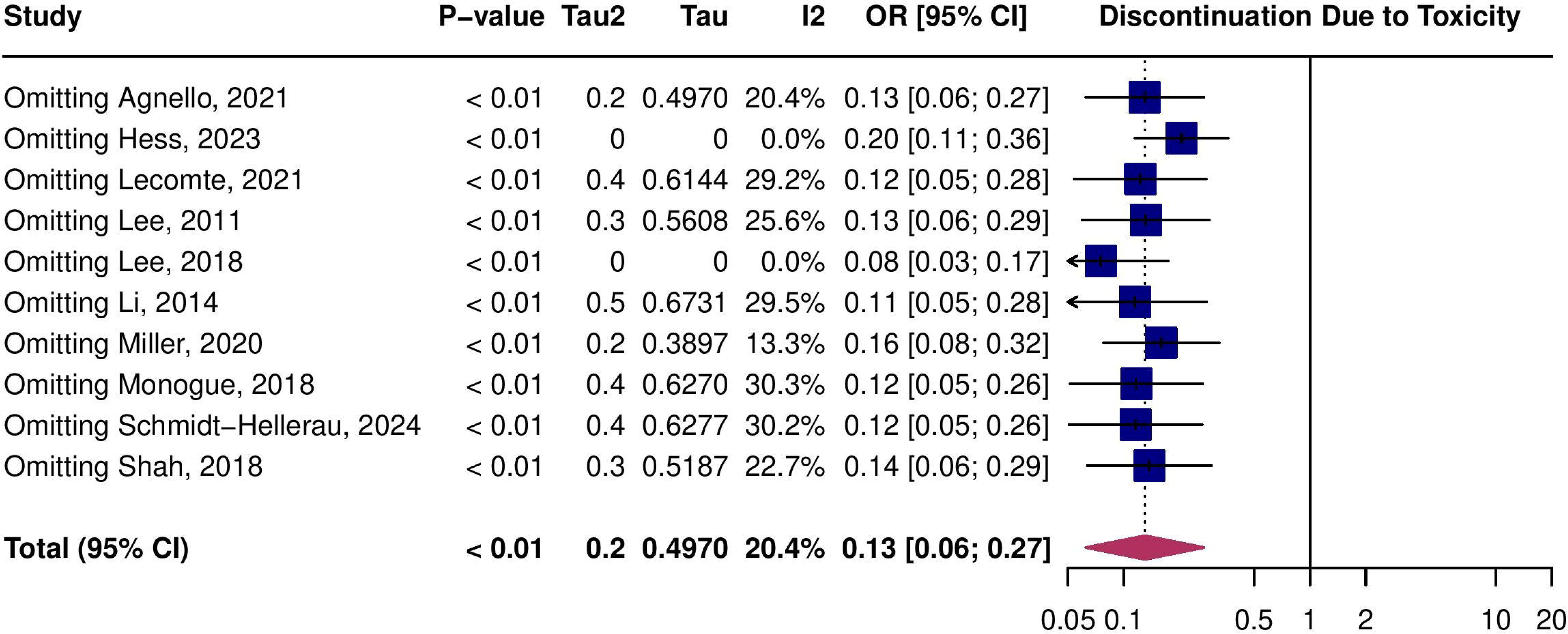
Leave-one-out meta-analysis of discontinuation due to toxicity for cefazolin vs. antistaphylococcal penicillins.

**Supplemental Figure 12.**
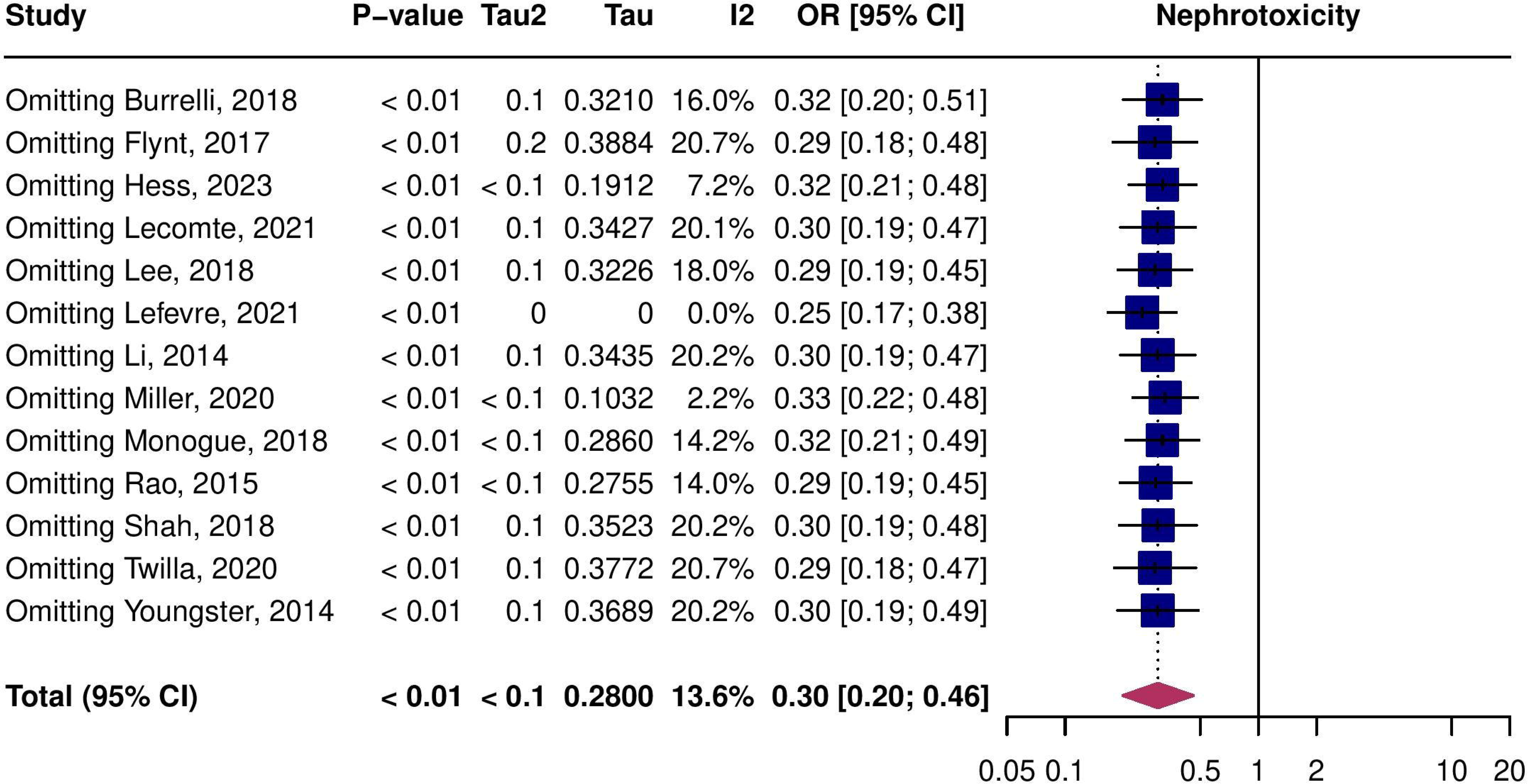
Leave-one-out meta-analysis of nephrotoxicity for cefazolin vs. antistaphylococcal penicillins.

## REFERENCES

1. Hindy J-R, Quintero-Martinez JA, Lee AT, et al. Incidence Trends and Epidemiology of Staphylococcus aureus Bacteremia: A Systematic Review of Population-Based Studies. Cureus 14:e25460.

2. Bai AD, Lo CKL, Komorowski AS, et al. *Staphylococcus aureus* bacteraemia mortality: a systematic review and meta-analysis. Clin Microbiol Infect 2022; 28:1076–1084.

3. Westgeest AC, Buis DTP, Sigaloff KCE, et al. Global Differences in the Management of Staphylococcus aureus Bacteremia: No International Standard of Care. Clin Infect Dis Off Publ Infect Dis Soc Am 2023; 77:1092–1101.

4. Lo CK-F, Sritharan A, Zhang J, et al. Clinical significance of cefazolin inoculum effect in serious MSSA infections: a systematic review. JAC-Antimicrob Resist 2024; 6:dlae069.

5. Delgado V, Ajmone Marsan N, de Waha S, et al. 2023 ESC Guidelines for the management of endocarditis: Developed by the task force on the management of endocarditis of the European Society of Cardiology (ESC) Endorsed by the European Association for Cardio-Thoracic Surgery (EACTS) and the European Association of Nuclear Medicine (EANM). Eur Heart J 2023; 44:3948–4042.

6. Baddour LM, Wilson WR, Bayer AS, et al. Infective Endocarditis in Adults: Diagnosis, Antimicrobial Therapy, and Management of Complications. Circulation 2015; 132:1435– 1486.

7. Gould FK, Denning DW, Elliott TSJ, et al. Guidelines for the diagnosis and antibiotic treatment of endocarditis in adults: a report of the Working Party of the British Society for Antimicrobial Chemotherapy. J Antimicrob Chemother 2012; 67:269–289.

8. McDonald EG, Aggrey G, Tarık Aslan A, et al. Guidelines for Diagnosis and Management of Infective Endocarditis in Adults: A WikiGuidelines Group Consensus Statement. JAMA Netw Open 2023; 6:e2326366.

9. Weis S, Kesselmeier M, Davis JS, et al. Cefazolin versus anti-staphylococcal penicillins for the treatment of patients with Staphylococcus aureus bacteraemia. Clin Microbiol Infect Off Publ Eur Soc Clin Microbiol Infect Dis 2019; 25:818–827.

10. Tong SYC, Mora J, Bowen AC, et al. The Staphylococcus aureus Network Adaptive Platform Trial Protocol: New Tools for an Old Foe. Clin Infect Dis Off Publ Infect Dis Soc Am 2022; 75:2027–2034.

11. Burdet C, Loubet P, Le Moing V, et al. Efficacy of cloxacillin versus cefazolin for methicillin-susceptible Staphylococcus aureus bacteraemia (CloCeBa): study protocol for a randomised, controlled, non-inferiority trial. BMJ Open 2018; 8:e023151.

12. Gravenkemper CF, Bennett JV, Brodie JL, Kirby WM. DICLOXACILLIN. IN VITRO AND PHARMACOLOGIC COMPARISONS WITH OXACILLIN AND CLOXACILLIN. Arch Intern Med 1965; 116:340–345.

13. Sidell S, Burdick RE, Brodie J, Bulger RJ, Kirby WM. New antistaphylococcal antibiotics. Comparative in vitro and in vivo activity of cephalothin, nafcillin, cloxacillin, oxacillin, and methicillin. Arch Intern Med 1963; 112:21–28.

14. Maraqa NF, Gomez MM, Rathore MH, Alvarez AM. Higher Occurrence of Hepatotoxicity and Rash in Patients Treated with Oxacillin, Compared with Those Treated with Nafcillin and Other Commonly Used Antimicrobials. Clin Infect Dis 2002; 34:50–54.

15. Viehman JA, Oleksiuk L-M, Sheridan KR, et al. Adverse Events Lead to Drug Discontinuation More Commonly among Patients Who Receive Nafcillin than among Those Who Receive Oxacillin. Antimicrob Agents Chemother 2016; 60:3090–3095.

16. Bidell MR, Patel N, O’Donnell JN. Optimal treatment of MSSA bacteraemias: a meta-analysis of cefazolin versus antistaphylococcal penicillins. J Antimicrob Chemother 2018; 73:2643–2651.

17. Rindone JP, Mellen CK. Meta-analysis of trials comparing cefazolin to antistaphylococcal penicillins in the treatment of methicillin-sensitive Staphylococcus aureus bacteraemia. Br J Clin Pharmacol 2018; 84:1258–1266.

18. Shi C, Xiao Y, Zhang Q, et al. Efficacy and safety of cefazolin versus antistaphylococcal penicillins for the treatment of methicillin-susceptible Staphylococcus aureus bacteremia: a systematic review and meta-analysis. BMC Infect Dis 2018; 18:508.

19. Lee BJ, Wang SK, Constantino-Corpuz JK, et al. Cefazolin vs. anti-staphylococcal penicillins for treatment of methicillin-susceptible Staphylococcus aureus bloodstream infections in acutely ill adult patients: Results of a systematic review and meta-analysis. Int J Antimicrob Agents 2019; 53:225–233.

20. Allen JM, Bakare L, Casapao AM, Klinker K, Childs-Kean LM, Pomputius AF. Cefazolin Versus Anti-Staphylococcal Penicillins for the Treatment of Patients with Methicillin-Susceptible Staphylococcus aureus Infection: A Meta-Analysis with Trial Sequential Analysis. Infect Dis Ther 2019; 8:671–686.

21. Page MJ, McKenzie JE, Bossuyt PM, et al. The PRISMA 2020 statement: an updated guideline for reporting systematic reviews. BMJ 2021; 372:n71.

22. Brooke BS, Schwartz TA, Pawlik TM. MOOSE Reporting Guidelines for Meta-analyses of Observational Studies. JAMA Surg 2021; 156:787–788.

23. Cochrane Handbook for Systematic Reviews of Interventions version 6.4. Cochrane, 2023. Available at: www.training.cochrane.org/handbook.

24. Veritas Health Innovation. Covidence systematic review software. Melbourne, Australia: Available at: www.covidence.org.

25. Sterne JA, Hernán MA, Reeves BC, et al. ROBINS-I: a tool for assessing risk of bias in non-randomised studies of interventions. BMJ 2016; 355:i4919.

26. RiskLofLbias VISualization (robvis): An R package and Shiny web app for visualizing riskLofLbias assessments - McGuinness - 2021 - Research Synthesis Methods - Wiley Online Library. Available at: https://onlinelibrary.wiley.com/doi/full/10.1002/jrsm.1411. Accessed 28 June 2024.

27. Viechtbauer W. Conducting Meta-Analyses in R with the metafor Package. J Stat Softw 8AD; 36:1–48.

28. Schwarzer G. meta: General Package for Meta-Analysis. 2022; Available at: https://CRAN.R-project.org/package=meta. Accessed 21 March 2022.

29. DerSimonian R, Laird N. Meta-analysis in clinical trials. Control Clin Trials 1986; 7:177–88.

30. Guyatt GH, Oxman AD, Vist GE, et al. GRADE: an emerging consensus on rating quality of evidence and strength of recommendations. BMJ 2008; 336:924–926.

31. Guideline Development Tool. Available at: https://gdt.gradepro.org/app/#projects/p_l_a_86476730-d11a-5706-b0c6-da0cc65f8a34_33ef0d3b-6762-4b0a-84c4-e6c7e2e2f643/evidence-syntheses/7429b174-6643-479c-b5d7-c59e84c4b699/quality-of-evidence. Accessed 21 October 2024.

32. B L, B H, F G, et al. Antistaphylococcal penicillins vs. cefazolin in the treatment of methicillin-susceptible Staphylococcus aureus infective endocarditis: a quasi-experimental monocentre study. Eur J Clin Microbiol Infect Dis Off Publ Eur Soc Clin Microbiol 2021; 40. Available at: https://pubmed.ncbi.nlm.nih.gov/34383175/. Accessed 15 October 2024.

33. Agnello S, Wardlow LC, Reed E, Smith JM, Coe K, Day SR. Clinical Outcomes of Daptomycin Versus Anti-Staphylococcal Beta-Lactams in Definitive Treatment of Methicillin-susceptible Staphylococcus aureus Bloodstream Infections. Int J Antimicrob Agents 2021; 58:106363.

34. Bai AD, Showler A, Burry L, et al. Comparative effectiveness of cefazolin versus cloxacillin as definitive antibiotic therapy for MSSA bacteraemia: results from a large multicentre cohort study. J Antimicrob Chemother 2015; 70:1539–1546.

35. Beganovic M, Cusumano JA, Lopes V, LaPlante KL, Caffrey AR. Comparative Effectiveness of Exclusive Exposure to Nafcillin or Oxacillin, Cefazolin, Piperacillin/Tazobactam, and Fluoroquinolones Among a National Cohort of Veterans With Methicillin-Susceptible Staphylococcus aureus Bloodstream Infection. Open Forum Infect Dis 2019; 6:ofz270.

36. Burrelli CC, Broadbent EK, Margulis A, et al. Does the Beta-Lactam Matter? Nafcillin versus Cefazolin for Methicillin-Susceptible Staphylococcus aureus Bloodstream Infections. Chemotherapy 2018; 63:345–351.

37. Js D, J T, S T. A large retrospective cohort study of cefazolin compared with flucloxacillin for methicillin-susceptible Staphylococcus aureus bacteraemia. Int J Antimicrob Agents 2018; 52. Available at: https://pubmed.ncbi.nlm.nih.gov/29499317/. Accessed 15 October 2024.

38. Al D, A M, M S, et al. Effectiveness and safety of cefazolin versus cloxacillin in endocarditis due to methicillin-susceptible Staphylococcus spp.: a multicenter propensity weighted cohort study. Eur J Clin Microbiol Infect Dis Off Publ Eur Soc Clin Microbiol 2024; 43. Available at: https://pubmed.ncbi.nlm.nih.gov/38806841/. Accessed 15 October 2024.

39. Flynt LK, Kenney RM, Zervos MJ, Davis SL. The Safety and Economic Impact of Cefazolin versus Nafcillin for the Treatment of Methicillin-Susceptible Staphylococcus aureus Bloodstream Infections. Infect Dis Ther 2017; 6:225–231.

40. Herrera-Hidalgo L, Muñoz P, Álvarez-Uría A, et al. Contemporary use of cefazolin for MSSA infective endocarditis: analysis of a national prospective cohort. Int J Infect Dis IJID Off Publ Int Soc Infect Dis 2023; 137:134–143.

41. Hess KA, Kooda K, Shulha JA, et al. Retrospective Evaluation of the Association of Oxacillin MIC on Acute Treatment Outcomes with Cefazolin and Antistaphylococcal Penicillins in Methicillin-Susceptible Staphylococcus aureus Bacteremia. J Clin Microbiol 2023; 61:e0003923.

42. Kimmig A, Weis S, Hagel S, Forstner C, Kesselmeier M, Pletz MW. Infektiologische Konsile bei Patienten mit Staphylococcus-aureus-Bakteriämie – eine retrospektive Beobachtungsstudie am Universitätsklinikum Jena. DMW - Dtsch Med Wochenschr 2018; 143:e179–e187.

43. Lee S, Song K-H, Jung S-I, et al. Comparative outcomes of cefazolin versus nafcillin for methicillin-susceptible Staphylococcus aureus bacteraemia: a prospective multicentre cohort study in Korea. Clin Microbiol Infect Off Publ Eur Soc Clin Microbiol Infect Dis 2018; 24:152–158.

44. Lecomte R, Bourreau A, Deschanvres C, et al. Comparative outcomes of cefazolin versus antistaphylococcal penicillins in methicillin-susceptible Staphylococcus aureus infective endocarditis: a post hoc analysis of a prospective multicentre French cohort study. Clin Microbiol Infect 2021; 27:1015–1021.

45. Lee S, Choe PG, Song K-H, et al. Is Cefazolin Inferior to Nafcillin for Treatment of Methicillin-Susceptible Staphylococcus aureus Bacteremia?L. Antimicrob Agents Chemother 2011; 55:5122–5126.

46. Li J, Echevarria KL, Hughes DW, Cadena JA, Bowling JE, Lewis JS. Comparison of Cefazolin versus Oxacillin for Treatment of Complicated Bacteremia Caused by Methicillin-Susceptible Staphylococcus aureus. Antimicrob Agents Chemother 2014; 58:5117–5124.

47. McDanel JS, Roghmann M-C, Perencevich EN, et al. Comparative Effectiveness of Cefazolin Versus Nafcillin or Oxacillin for Treatment of Methicillin-Susceptible Staphylococcus aureus Infections Complicated by Bacteremia: A Nationwide Cohort Study. Clin Infect Dis Off Publ Infect Dis Soc Am 2017; 65:100–106.

48. Miller MA, Fish DN, Barber GR, et al. A comparison of safety and outcomes with cefazolin versus nafcillin for methicillin-susceptible Staphylococcus aureus bloodstream infections. J Microbiol Immunol Infect Wei Mian Yu Gan Ran Za Zhi 2020; 53:321–327.

49. Ht M, Cb T, S B, et al. Treatment outcomes with benzylpenicillin and non-benzylpenicillin antibiotics, and the performance of the penicillin zone-edge test versus molecular detection of blaZ in penicillin-susceptible Staphylococcus aureus (PSSA) bacteraemia. J Antimicrob Chemother 2023; 78. Available at: https://pubmed.ncbi.nlm.nih.gov/37596905/. Accessed 15 October 2024.

50. Monogue ML, Ortwine JK, Wei W, Eljaaly K, Bhavan KP. Nafcillin versus cefazolin for the treatment of methicillin-susceptible Staphylococcus aureus bacteremia. J Infect Public Health 2018; 11:727–731.

51. Paul M, Zemer-Wassercug N, Talker O, et al. Are all beta-lactams similarly effective in the treatment of methicillin-sensitive Staphylococcus aureus bacteraemia? Clin Microbiol Infect Off Publ Eur Soc Clin Microbiol Infect Dis 2011; 17:1581–1586.

52. Pollett S, Baxi SM, Rutherford GW, Doernberg SB, Bacchetti P, Chambers HF. Cefazolin versus Nafcillin for Methicillin-Sensitive Staphylococcus aureus Bloodstream Infection in a California Tertiary Medical Center. Antimicrob Agents Chemother 2016; 60:4684–4689.

53. Sn R, Nj R, Bj L, et al. Treatment outcomes with cefazolin versus oxacillin for deep-seated methicillin-susceptible Staphylococcus aureus bloodstream infections. Antimicrob Agents Chemother 2015; 59. Available at: https://pubmed.ncbi.nlm.nih.gov/26077253/. Accessed 15 October 2024.

54. Renaud CJ, Lin X, Subramanian S, Fisher DA. High-dose cefazolin on consecutive hemodialysis in anuric patients with Staphylococcal bacteremia. Hemodial Int Int Symp Home Hemodial 2011; 15:63–68.

55. Reynolds G, Crawford S, Cuenca J, Ghosh N, Newton P. Penicillin versus anti-staphylococcal beta-lactams for penicillin-susceptible Staphylococcus aureus blood stream infections: a retrospective cohort study. Eur J Clin Microbiol Infect Dis Off Publ Eur Soc Clin Microbiol 2022; 41:147–151.

56. Schmidt-Hellerau K, Breuninger M, Kessel J, et al. Flucloxacillin and cefazolin for treatment of Staphylococcus aureus bloodstream infection. Infection 2024; 52:1159–1163.

57. Shah MD, Wardlow LC, Stevenson KB, Coe KE, Reed EE. Clinical Outcomes with Penicillin Versus Alternative β-Lactams in the Treatment of Penicillin-Susceptible Staphylococcus aureus Bacteremia. Pharmacotherapy 2018; 38:769–775.

58. Twilla JD, Algrim A, Adams EH, Samarin M, Cummings C, Finch CK. Comparison of Nafcillin and Cefazolin for the Treatment of Methicillin-Susceptible Staphylococcus aureus Bacteremia. Am J Med Sci 2020; 360:35–41.

59. Willekens R, Puig-Asensio M, Suanzes P, et al. Empirical use of β-lactam/β-lactamase inhibitor combinations does not increase mortality compared with cloxacillin and cefazolin in methicillin-susceptible Staphylococcus aureus bacteraemia: a propensity-weighted cohort study. J Antimicrob Chemother 2022; 77:2288–2295.

60. Za Y, Rb K, Jr G, S C, Om AS. Post-treatment outcomes of ceftriaxone versus antistaphylococcal penicillins or cefazolin for definitive therapy of methicillin-susceptible Staphylococcus aureus bacteremia. Eur J Clin Microbiol Infect Dis Off Publ Eur Soc Clin Microbiol 2023; 42. Available at: https://pubmed.ncbi.nlm.nih.gov/36800065/. Accessed 15 October 2024.

61. Youngster I, Shenoy ES, Hooper DC, Nelson SB. Comparative evaluation of the tolerability of cefazolin and nafcillin for treatment of methicillin-susceptible Staphylococcus aureus infections in the outpatient setting. Clin Infect Dis Off Publ Infect Dis Soc Am 2014; 59:369–375.

62. Lefèvre B, Hoen B, Goehringer F, et al. Antistaphylococcal penicillins vs. cefazolin in the treatment of methicillin-susceptible Staphylococcus aureus infective endocarditis: a quasi-experimental monocentre study. Eur J Clin Microbiol Infect Dis 2021; 40:2605–2616.

63. Duke C, Parker SL, Zam BB, et al. Population pharmacokinetics of unbound cefazolin in infected hospitalized patients requiring intermittent high-flux haemodialysis: can a three-times-weekly post-dialysis dosing regimen provide optimal treatment? J Antimicrob Chemother 2024; :dkae318.

