## Supplementary material for "Cefazolin versus Antistaphylococcal Penicillins for the Treatment of Methicillin-Susceptible *Staphylococcus aureus* Bacteremia: A Systematic Review and Meta-Analysis": Tables and Supplemental Tables

**Supplemental Table 1.** Summary of first-line recommended backbone therapy for methicillin-sensitive native valve *S. aureus* IE from different guidelines.

| Guideline, Year | First-Line Agent for Methicillin-Sensitive SAB | Strength of Recommendation |
| --- | --- | --- |
| European Society of Cardiology, 2023 | Cefazolin 2g IV q8h or (flu)cloxacillin 12g IV daily in 4-6 doses | IB |
| American Heart Association Guidelines, 2015 | Nafcillin or oxacillin 12g IV daily in 4-6 doses | IC |
| British Society for Antimicrobial Chemotherapy, 2012 | Flucloxacillin 2g IV q6h | A |
| WikiGuidelines, 2023 | Cefazolin 2g IV q8h or (flu)cloxacillin, nafcillin, or oxacillin 2g IV q4h | NR |

NR=Not reported

**Supplemental Table 2.** Detailed search strategy (updated from Weis et al.)

| **Database** | **Search Terms** | **Hits** |
| --- | --- | --- |
| PubMed | (((((((((((((((flucloxacillin) OR nafcillin) OR methicillin) OR dicloxacillin) OR oxacillin) OR floxacillin) OR cloxacillin) OR betalactam) OR betala*) OR penicillin) OR beta-la*) OR isoxazolylpenicillin)) AND (((((((((blood stream) OR blood) OR septicemia) OR septicemia) OR bacteremia) OR bacteraemia) OR bacteraemia) OR Blutstrom*) OR bakteriamie)) AND (((staphylococcus aureus[MeSH Terms]) OR aureus) OR staphylococcus aureus)) AND (((cefazolin) OR cefazolin) OR cephazolin) AND ("2018/06/26"[Date - Publication] : "2024/08/29"[Date - Publication]) | 122 |
| Web of Science | (((((((((((((((flucloxacillin) OR nafcillin) OR methicillin) OR dicloxacillin) OR oxacillin) OR fluoxacillin) OR cloxacillin) OR betalactams) OR betala*) OR penicillin) OR beta-la*) OR isoxazolylpenicillins)) AND (((((((((blood stream) OR blood) OR septicemia) OR septicemia) OR bacteremia) OR bacteraemia) OR bacteraemia) OR Blutstrom*) OR Bakteriämie)) AND (((staphylococcus aureus[MeSH Terms]) OR aureus) OR staphylococcus aureus)) AND (((cefazolin) OR Cefazolin) OR cephazoline) (All Fields) and 2018-2024 (Year Published) | 172 |
| Cochrane Database of Systemic Reviews | ((aureus OR staphylococcus aureus) or MeSH descriptor: [Staphylococcus aureus] explode all trees) and ((blood stream OR (bacteremia) OR (bacteraemia)) or MeSH descriptor: [Bacteremia] explode all trees) and (Cefazolin OR Cephazolin OR beta-lactam OR anti-staphylo* OR nafcillin OR dicloxacillin OR flucloxacillin OR cloxacillin) | 18 |
| Clinicaltrials.gov | (Cefazolin) AND Staphylococcus Aureus AND Status completed/terminated | 3 |

**Supplemental Table 3.** Study characteristics.

| Study | Country | Number of Centers | Study Design | Total N Cefazolin | Total N ASPs | ASP Type | Cefazolin Percent with IE | ASP Percent with IE | Cefazolin Percent Admitted to ICU | ASP Percent Admitted to ICU |
| --- | --- | --- | --- | --- | --- | --- | --- | --- | --- | --- |
| Agnello, 2021 | USA | 1 | Retrospective cohort | 30 | 30 | Nafcillin | 6.7 | 16.7 | 20.0 | 16.7 |
| Bai, 2015 | Canada | 6 | Retrospective cohort | 105 | 249 | Cloxacillin | 1.9 | 12.0 | 9.5 | 18.1 |
| Beganovic, 2019 | USA | NR | Retrospective cohort | 44 | 44 | Not specified | NR | NR | NR | NR |
| Burrelli, 2018 | USA | 1 | Retrospective cohort | 41 | 116 | Nafcillin | 9.8 | 12.1 | 36.6 | 25.9 |
| Davis, 2018 | Australia, New Zealand | 27 | Prospective cohort | 792 | 6520 | Flucloxacillin | 5.9 | 8.0 | NR | NR |
| Destrem, 2024* | France | 6 | Retrospective cohort | 98 | 94 | Cloxacillin | 100.0 | 100.0 | NR | NR |
| Flynt, 2017 | USA | 4 | Retrospective cohort | 68 | 81 | Nafcillin | 16.2 | 27.2 | NR | NR |
| Herrera-Hidalgo | Spain | 39 | Prospective cohort | 57 | 537 | Cloxacillin | 100.0 | 100.0 | NR | NR |
| Hess, 2023 | USA | 1 | Retrospective cohort | 235 | 167 | Nafcillin, Oxacillin | 10.6 | NR | NR | NR |
| Kimmig, 2018 | Germany | 1 | Retrospective cohort | 61 | 131 | Flucloxacillin | NR | NR | NR | NR |
| Lecomte, 2021 | France | 2 | Prospective cohort | 53 | 157 | Not specified | 100.0 | 100.0 | 20.8 | 38.2 |
| Lee, 2011 | South Korea | 1 | Retrospective cohort | 41 | 41 | Nafcillin | 2.4 | 2.4 | NR | NR |
| Lee, 2018 | South Korea | 10 | Prospective cohort | 79 | 163 | Nafcillin | 1.3 | 6.7 | NR | NR |
| Lefevre, 2021 | France | 1 | Retrospective cohort | 38 | 35 | Not specified | 100.0 | 100.0 | NR | NR |
| Li, 2014 | USA | 2 | Retrospective cohort | 59 | 34 | Oxacillin | 23.7 | 8.8 | 6.8 | 17.6 |
| McDanel, 2017 | USA | 119 | Retrospective cohort | 1163 | 2004 | Not specified | 4.5 | 7.2 | 15.3 | 18.9 |
| Miller, 2020 | USA | 1 | Retrospective cohort | 51 | 79 | Nafcillin | 13.7 | 19.0 | 43.1 | 55.7 |
| Mok, 2023 | Singapore | 1 | Retrospective cohort | 10 | 27 | Cloxacillin | NR | NR | NR | NR |
| Monogue, 2018 | USA | 1 | Retrospective cohort | 71 | 71 | Nafcillin | 4.2 | 8.5 | 11.3 | 32.4 |
| Paul, 2011 | Israel | 1 | Retrospective cohort | 72 | 281 | Cloxacillin | NR | NR | NR | NR |
| Pollett, 2016 | USA | 1 | Retrospective cohort | 70 | 30 | Nafcillin | 14.3 | 16.7 | 12.9 | 26.7 |
| Rao, 2015 | USA | 2 | Retrospective cohort | 103 | 58 | Oxacillin | 16.5 | 20.7 | 41.7 | 32.8 |
| Renaud, 2011 | Singapore | 1 | Prospective cohort | 14 | 13 | Cloxacillin | 0.0 | 7.7 | NR | NR |
| Reynolds, 2022 | Australia | NR | Retrospective cohort | 12 | 14 | Flucloxacillin | NR | NR | 33.3 | 21.4 |
| Schmidt-Hellerau, 2024 | Germany | 2 | Prospective cohort | 15 | 56 | Flucloxacillin | 20.0 | 8.9 | NR | NR |
| Shah, 2018 | USA | 1 | Retrospective cohort | 35 | 45 | Nafcillin | NR | NR | NR | NR |
| Twilla, 2020 | USA | 4 | Retrospective cohort | 151 | 126 | Nafcillin | 8.6 | 15.9 | NR | NR |
| Willekens, 2022 | Spain | 1 | Prospective cohort | 14 | 57 | Cloxacillin | NR | NR | NR | NR |
| Yetmar, 2023 | USA | 5 | Retrospective cohort | 168 | 18 | Nafcillin, Oxacillin | NR | NR | NR | NR |
| Youngster, 2014 | USA | 1 | Retrospective cohort | 119 | 366 | Nafcillin | 5.9 | 7.7 | NR | NR |

NR=Not reported

*91.1% were MSSA, 8.9% were coagulase-negative Staphylococci

**Supplemental Table 4.** Study definitions of treatment assignment, TRAEs, and nephrotoxicity.

| **Study** | **Treatment Assignment** | **TRAEs** | **Nephrotoxicity** |
| --- | --- | --- | --- |
| Agnello, 2021 | Definitive therapy, defined as the antibiotic administered for at least 14 days after susceptibilities were resulted or the antibiotic prescribed on hospital discharge | NA | NA |
| Bai, 2015 | Antibiotic used for >3 days and >50% of the duration | NA | NA |
| Beganovic, 2019 | The only antibiotic administered for the duration of treatment. | NA | NA |
| Burrelli, 2018 | Antibiotic received for >72 hours as directed therapy and <72 hours of any empiric antibiotics | Acute kidney injury, acute interstitial nephritis, or hepatotoxicity within 30 days of treatment cessation | Creatinine increase of 44.2 umol/L or 50% from baseline during hospitalization |
| Davis, 2018 | Definitive antibiotic, defined as the antibiotic used once the susceptibility of the isolate was known | NA | NA |
| Destrem, 2024 | First antibiotic received for >10 consecutive days | NA | Increase of baseline creatinine by 50% at day 14 |
| Flynt, 2017 | The first antibiotic among cefazolin or nafcillin received for >72 hours | C. difficile infection, neutropenia, transaminitis, or rash | Increase in baseline creatinine by 26.4umol/L in 48 hours or by 50% |
| Herrera-Hidalgo, 2023 | Antibiotic received for >75% of the treatment length | NR | NA |
| Hess, 2023 | Definitive therapy, defined as the first antibiotic given for >48 hours once susceptibilities were known. | NR | NR |
| Kimmig, 2018 | NR | NA | NA |
| Lecomte, 2021 | Receipt of at least one dose of cefazolin or an antistaphylococcal penicillin (but not both) | NA | NR |
| Lee, 2011 | Cefazolin was used during a period of nafcillin unavailability, although patients with suspected central nervous system infections were treated with other antibiotics. Outside the period of unavailability, nafcillin was the preferred agent. | NA | NA |
| Lee, 2018 | Definitive antibiotic, defined as the antibiotic received after susceptibility was resulted | NA | NR |
| Lefevre, 2021 | Assignment was based on availability of the antibiotics. Antistaphylococcal penicillins were used initially, but after a shortage cefazolin was favored. Patients who received both were excluded. | Transaminitis/bilirubin elevation or acute kidney injury | Elevation of 1.5 times the baseline or an increase in 26.5umol/L |
| Li, 2014 | >10 days of the respective antibiotic | Adverse events while on therapy | Increase in baseline creatinine of 44.2umol/L or by 50% |
| McDanel, 2017 | Definitive therapy, defined as the antibiotic received between days 4 and 14 after the first positive blood culture was collected | NA | NA |
| Miller, 2020 | Receipt of >24h of the respective antibiotic | Acute kidney injury, thrombocytopenia, transaminitis or elevated bilirubin, allergic reactions not attributable to another cause and/or change or interruption in the treatment due to documented intolerance/adverse event | Increase in baseline creatinine be 44.2umol/L or by 50% |
| Mok, 2023 | Definitive therapy, defined as the antibiotic given after 3 days of the collection of the first positive blood culture | Adverse events attributed to the antibiotic | NR |
| Monogue, 2018 | Receipt of the respective antibiotic for >72 hours | Nephrotoxicity, neutropenia, thrombocytopenia drug-induced fever, or infusion site reaction | Increase in baseline creatinine of 44.2umol/L or by 50% |
| Paul, 2011 | Definitive treatment, defined as the antibiotic received on days 3-9 after collection of the positive blood culture | NA | NA |
| Pollett, 2016 | Receipt of the respective antibiotic for >5 days within <7 days of blood culture positivity and excluding patients that received both cefazolin and nafcillin | NA | NA |
| Rao, 2015 | Definitive treatment within <48 hours of blood culture positivity | NR | Increase in baseline creatinine of 44.2umol/L or by 50% during hospitalization |
| Renaud, 2011 | Before and after the implementation of a cefazolin protocol for MSSA bacteremia in patients on hemodialysis, where previously cloxacillin was used | NR | NA |
| Reynolds, 2022 | Definitive treatment, defined as the antibiotic received for the majority of the course of therapy | NA | KDIGO classification |
| Schmidt-Hellerau, 2024 | Initial therapy for MSSA bacteremia | NA | NR |
| Shah, 2018 | Definitive treatment, defined as the antibiotic used after susceptibilities were available | Rash, gastrointestinal intolerance, and nephrotoxicity | Increase in baseline creatinine of 44.2umol/L or by 50% |
| Twilla, 2020 | Antibiotic received for >3 days and for >50% of the duration of therapy | Acute kidney injury, transaminitis, C. difficile infection, neutropenia, and rash | NR |
| Willekens, 2022 | Antibiotic that was started initially | NA | NA |
| Yetmar, 2023 | Antibiotic used for the majority of outpatient treatment | NA | NA |
| Youngster, 2014 | Antibiotic prescribed for outpatient parenteral antimicrobial therapy | Rash, nephrotoxicity, transaminitis, neutropenia, thrombocytopenia, eosinophilia, and C. difficile infection | Increase in baseline creatinine of 44.2umol/L or by 50% |

**Supplemental Table 5.** GRADE assessment.

| **Certainty assessment** | | | | | | | **№ of patients** | | **Effect** | | **Certainty** | **Importance** |
| --- | --- | --- | --- | --- | --- | --- | --- | --- | --- | --- | --- | --- |
| **№ of studies** | **Study design** | **Risk of bias** | **Inconsistency** | **Indirectness** | **Imprecision** | **Other considerations** | **[intervention]** | **[comparison]** | **Relative (95% CI)** | **Absolute (95% CI)** |  |  |
| **30-Day Mortality** | | | | | | | | | | | | |
| 16 | non-randomised studies | very serious | not serious | not serious | not serious | none | 227/2631 (8.6%) | 1113/9338 (11.9%) | **OR 0.73** (0.62 to 0.85) | **29 fewer per 1,000** (from 42 fewer to 16 fewer) | ⨁⨁◯◯ Low | CRITICAL |
| **90-Day Mortality** | | | | | | | | | | | | |
| 14 | non-randomised studies | very serious | not serious | not serious | serious | none | 359/2097 (17.1%) | 801/3318 (24.1%) | **OR 0.80** (0.61 to 1.05) | **38 fewer per 1,000** (from 79 fewer to 9 more) | ⨁◯◯◯ Very low | CRITICAL |
| **TRAE** | | | | | | | | | | | | |
| 14 | non-randomised studies | very serious | serious | not serious | not serious | strong association | 116/1052 (11.0%) | 412/1755 (23.5%) | **OR 0.33** (0.18 to 0.63) | **143 fewer per 1,000** (from 182 fewer to 73 fewer) | ⨁⨁◯◯ Low | IMPORTANT |
| **Discontinuation Due to Toxicity** | | | | | | | | | | | | |
| 10 | non-randomised studies | very serious | not serious | not serious | not serious | very strong association | 14/669 (2.1%) | 142/843 (16.8%) | **OR 0.13** (0.06 to 0.27) | **143 fewer per 1,000** (from 156 fewer to 117 fewer) | ⨁⨁⨁⨁ High | CRITICAL |
| **Nephrotoxicity** | | | | | | | | | | | | |
| 13 | non-randomised studies | very serious | not serious | not serious | not serious | strong association | 48/1103 (4.4%) | 205/1498 (13.7%) | **OR 0.30** (0.20 to 0.46) | **91 fewer per 1,000** (from 106 fewer to 69 fewer) | ⨁⨁⨁◯ Moderate | IMPORTANT |
